# Trajectories of pain and cognitive function: 22 years of evidence in mid-to-later life

**DOI:** 10.64898/2026.02.10.26345971

**Authors:** Salomé Andres, Simon R Cox, Chloe Fawns-Ritchie

## Abstract

Chronic pain has been identified as a risk factor for cognitive decline in later life. However, most studies measure pain at a single time point and none have investigated whether variations in pain severity are associated with changes in cognitive function over time. This project aimed to assess the relationship between individual-level change in pain severity and decline in cognitive function over time. We used data from the English Longitudinal Study of Ageing (ELSA), a cohort of nationally representative middle aged and older adults. Pain severity was measured at each wave using a 4-point scale (none, mild, moderate and severe) and cognitive function was assessed using 3 objective tests. We applied latent growth curve modelling, a method for longitudinal analysis, to 19,376 ELSA participants’ data collected over 11 waves, spanning more than 20 years, to examine the relationship between initial level and change of both pain and cognitive function.

Adjusting for age and sex, worsening chronic pain severity was associated with accelerated decline in a general measure of cognitive function (β = −0.053, *p* = 0.039). However, when additionally adjusting for ethnicity, socioeconomic status and comorbid chronic conditions, this association was attenuated to non-significance (β = −0.025, *p* = 0.365). Greater initial pain severity was associated with steeper decline in cognitive function even in the fully adjusted model (β = −0.104, *p* < 0.001). Our study suggests that baseline level of pain severity but not worsening pain severity is associated with steeper decline in cognitive function over time.

**SUMMARY:** Age- and sex-adjusted analyses find that higher baseline and worsening pain severity predict faster cognitive decline; only baseline pain remains significant after full adjustment.

## INTRODUCTION

Chronic pain and cognitive decline represent two major public health challenges, particularly in ageing populations. Chronic pain affects an estimated 35-51% of adults in the UK, rising to over 60% in adults aged over 75 years.[14] Concurrently, key cognitive functions such as memory and processing speed decline with increasing age,[48,51] though rates of decline vary between individuals.[60,53,11,19] Identifying potential risk factors of accelerated cognitive decline is crucial and chronic pain’s contribution has not been wholly explored.

Individuals with chronic pain tend to perform worse on various cognitive tests, including memory, executive function and processing speed, compared to those without pain.[31,7,8,62,21,28] This cross-sectional evidence cannot distinguish between stable between-person differences and within-person changes in cognitive function over time, nor establish the temporal ordering of pain and cognition. Longitudinal findings are mixed, with some studies reporting that presence of pain, greater pain severity, or more years of pain duration are associated with steeper cognitive decline or incidence of cognitive impairment[5,44,57]. However, a recent meta-analysis[1] of 10 longitudinal studies (57,495 older adults) reported that persistent pain at baseline was not associated with incident cognitive decline up to 11.8 years later (RR = 1.05; 95% CI 0.92 to 1.21).

Although these longitudinal studies measure cognitive function longitudinally, most evaluate pain only once, despite evidence of the temporal instability of pain presence and severity.[12,13,16,27] Cross-sectional designs are particularly vulnerable to measurement error and confounding (reverse causation, time-invariant confounding, dynamic exposure misclassification), particularly in the context of a subjective measure such as pain which is standardised internally by participants. Longitudinal studies, by capturing these processes dynamically, minimise these confounds and improve reliability, inferential precision, and power.[40,41] As pain may emerge, intensify or remit during follow-up, modelling longitudinal change in *both* pain and cognitive function over time, offers a more stringent test of a causal hypothesis with correlational data. Milani et al.[30] analysed two waves of data over 4 years (n = 2,349) using cross-lagged models, and found that higher-than-expected pain interference (i.e. whether pain interfered with daily activity) at follow-up was associated with lower-than-expected scores on a brief cognitive screening test, after controlling for baseline scores (β = −0.07, p < 0.01). Another study[20] approached this question using group-based trajectory modelling which categorised participants in 3 distinct groups (low-stable, high-stable, and moderate-increasing) based on their changes in the probability of presence or absence of pain across time rather than fluctuations in pain severity. Using linear mixed-effect models, the authors found that the moderate-increasing group exhibited steep decline on executive function (β = −0.005 SD/year, p = .012) and global cognition (β = −0.014 SD/year, p < .001); however, this method assumes all group members share the same trajectory, neglecting within-person variation.

The current study addresses existing gaps by applying latent growth curve modelling (LGCM) and leveraging 22 years of data collected on both pain severity and cognitive function from 19,376 community-dwelling adults. This approach allows for the investigation of the effect of *within-person* changes in pain severity over time on concurrent *within-person* changes in cognitive function.

## METHODS

### Study population

This study leverages data from the English Longitudinal Study of Ageing (ELSA),[3,52] a population-based cohort study designed to be representative of community-dwelling adults aged 50 years and older residing in England. Data collection commenced in 2002 (wave 1 n = 12,099) and participants were recruited from responders to the Health Survey for England[33] over the age of 50 and their spouse. Participants have been followed up approximately every two years, and the sample has been refreshed at waves 3, 4, 6, 7, 9, 10 and 11 to ensure representativeness is maintained. To date, 11 waves of data have been collected from a total of 23,259 participants. Since the present study aims to investigate cognitive decline in mid-to-later life, we used data from the 19,376 participants who were over the age of 40 in 2002 which marks the start of data collection for wave 1.

Data were collected via face-to-face interviews conducted in the participant’s own homes and via self-completion questionnaires. Information collected included sociodemographic, health and lifestyle reports. Although all participants were recruited whilst they resided in the community, a small portion of subsequent interviews took place in institutions (0.6% in our sample) if the participant moved to a care home or similar establishment. Detailed descriptions of the study design and data collected are reported elsewhere.[61]

Data were downloaded from the UK Data Service in May 2025. ELSA has received ethical approval for all waves of data collection (most recently from South Central – Berkshire Research Ethics Committee on 22^nd^ May 2023; REF: 23/SC/0112).

### Measures

#### Pain severity

At each wave, participants were asked “Are you often troubled with pain?” Participants who answered “yes” were then asked “How bad is the pain most of the time? Is it mild, moderate, or, severe?”. To capture pain severity in the current study, we created a four-point ordinal scale (none, mild, moderate, severe) from these answers.

#### Cognitive function

The current study used scores on 3 of the cognitive tests. Memory was assessed with a recall test (word-list learning): Participants heard a list of 10 words that they had to recall immediately (immediate recall) and after a short delay (delayed recall), during which they completed other cognitive tests. Scores on the immediate and delayed recall tests were strongly correlated (*r* = 0.80, *p* < 0.05) and combined into a word recall variable by summing these results (range 0 to 20). Data for the recall test were available at all 11 waves. Executive function was assessed with animal naming (word-finding), a category fluency test where participants were asked to name as many animals as possible in 60 seconds. The score was the number of unique animals named within the time limit. Animal naming was collected at all waves, except wave 6. Finally, processing speed was assessed with letter cancellation, which involved participants crossing out as many Ps and Ws on a sheet of paper as fast as possible in 60 seconds. The score was the number of letters correctly scored out within the time limit. Letter cancellation was only assessed at waves 1 to 5.

Scores of 0 in animal naming (n = 727) and letter cancellation (n = 28) were set to missing as these likely indicate the participant did not complete or understand the task. After visual inspection of the distribution of all three cognitive tests (Supplementary I), scores 4 standard deviations above the mean for animal naming (4SD = 54; n = 21) were set to missing as these results are extremely high given the time limit, and potentially suggest an administration or data entry error. There were no outliers in the recall tests plots and the letter cancellation appeared to have a heavy-tailed distribution rather than isolated anomalies so no further alterations were made to either variable.

#### Covariates

Age, sex, ethnicity, socioeconomic status and comorbidities were used as covariates. Unless otherwise specified, covariate data were taken from the first wave in which participants provided data. Age was measured in years and participants under 40 in 2002 (at wave 1) were removed from the dataset (n = 3,883). To avoid disclosure due to small numbers, ELSA set the age of all participants over 90 years to a value 99. Here, we recoded these individuals’ ages to 91 years. To improve precision of the age variable, we used information from ELSA wave 0 (i.e., the Health Survey for England 1998-2001) for participants whose age was recorded at 99 in wave 1 (n = 62) and added 4 years (time elapsed between waves 0 and 1) to their reported age at wave 0. Finally, the age variable was standardised (mean = 0; SD = 1) before being added to the model to help convergence. Sex was recorded as a binary, male or female. The ethnicity variable was collapsed by ELSA to “white” and “non-white” to avoid disclosure. Socioeconomic status was based on the 8-point scale of the National Statistics Socio-Economic Classification ranging from “higher managerial and professional occupations” to “never worked or long-term unemployed”.[35] Comorbidity variables were derived from responses to the question “has a doctor ever told you that you have (or have had) [list of medical conditions]”. These include diabetes (or high blood sugar), hypertension (high blood pressure), heart disease (angina, a heart attack, congestive heart failure, an abnormal heart rhythm or any other heart trouble), lung disease, stroke and cancer or a malignant tumour (excluding minor skin cancers). Participants were classified as having a specific health condition if they ever reported it, at any wave of data collection.

### Statistical analysis

To estimate longitudinal trajectories of pain and cognitive function, we used latent growth curve modelling (LGCM), a structural equation modelling (SEM) technique which estimates both individual- and group-level trajectories over time. Conceptually, SEM allows researchers to investigate variables which are unobservable and measured indirectly by multiple indicators. LGCM explicitly parses the longitudinal data into an initial starting point (a latent intercept) and change over time (a latent slope).[25] This specific framework allows for the relationship between intercepts and slopes of both pain severity and cognitive decline to be estimated and their associations tested within the same simultaneous model.

Using observed pain severity scores across 11 waves, we derived two latent variables which capture the intercept (essentially pain severity level at baseline) and slope (linear rate of change in pain severity across the 11 waves) for each participant. To convert the four observed ordinal values to a continuous latent variable, the model determined three thresholds, which are the cut-off points on the underlying latent variable that that mark where the probability of endorsing one ordinal category shifts to the next.

To model cognitive function we used the hierarchical “factor-of-curves”[29,59] method: for each individual cognitive test (word recall, animal naming and letter cancellation), we modelled a latent intercept (cognitive level at baseline) and slope (rate of change in the cognitive test across the 11 waves) using the observed cognitive test scores across waves. We imposed a superordinate latent measure of general cognitive function (‘*g*’; top half of Figure 1) to reflect the well-known correlational structure across cognitive tests and their correlated changes over time[10,55]. To allow identification, the marker variable approach was employed, where the loading of the word recall latent intercept and slope on higher-order factors was fixed at 1 and the mean intercept and slope of recall set to 0. The slope loadings for each cognitive test reflected the mean time in years since wave one (0, 2.29, 4.29, 6.25, 8.29, 10.21, 12.21, 14.21, 16.29, 19.79, and 21.59, estimated from the month and year of data collection at each wave), fixing the slope estimated scale to change in standard deviation units per year.

**Figure 1:**
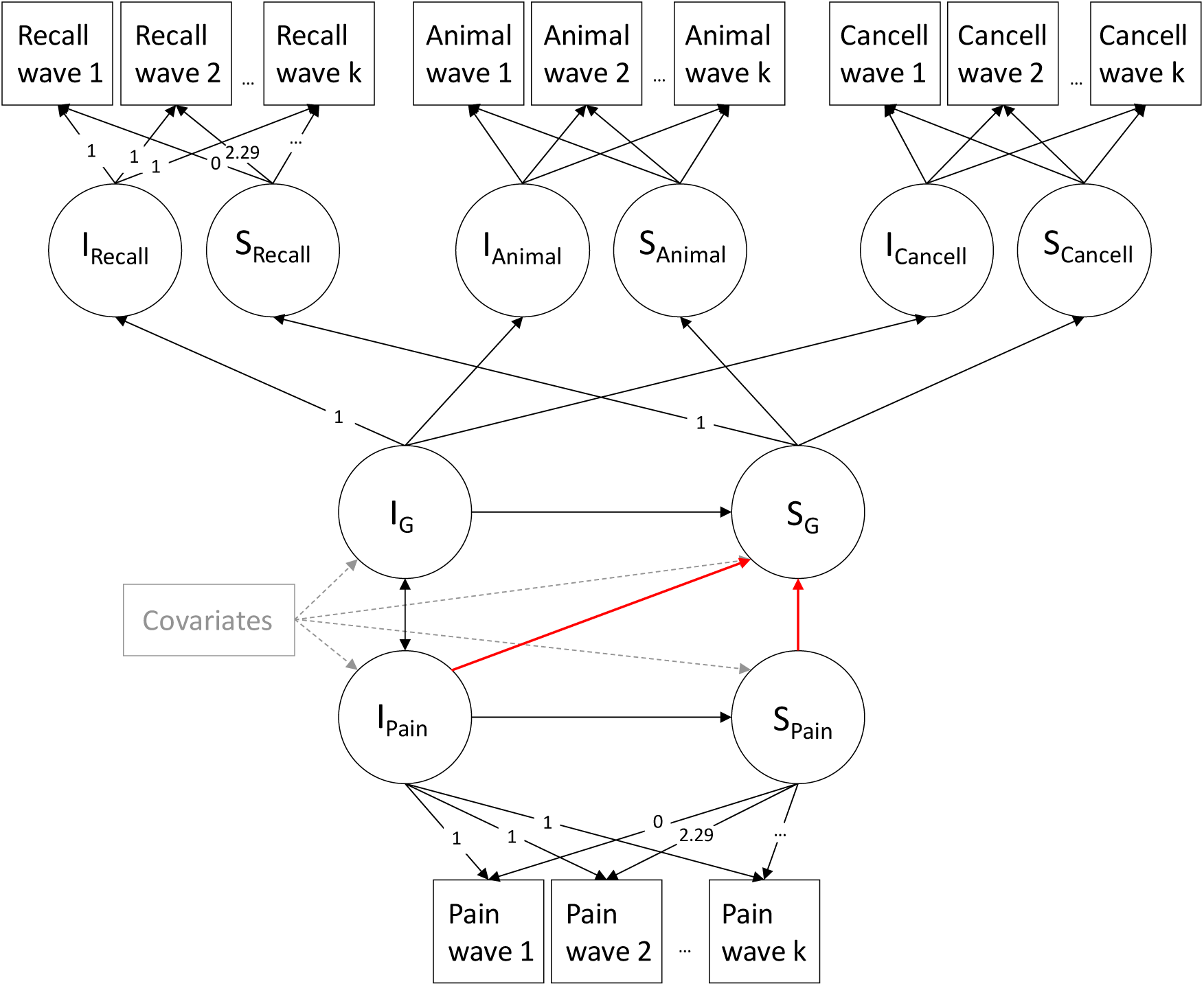
Simplified path diagram of the parallel process latent growth curve model of pain severity and general cognitive function. Circles are latent factors, squares represent observed variables, single-headed arrows are regression paths and double-headed arrows are correlation paths. The red arrows are the focus of our hypothesis. Dashed grey lines are regression paths for covariates. I = Intercept, S = Slope, G = general cognitive function, Animal = Animal naming test, Cancell = Letter cancellation test, Recall = Word recall test. For illustrative purposes, the loadings for animal naming and letter cancellation are not displayed; however, these are the same as the loading for recall and pain. Following the marker variable approach, the word recall test’s loadings on general cognitive function intercept and slopes was set to 1 and the means of intercept and slope of the word recall test were set to 0.

Model fit was assessed using root-mean-square error of approximation (RMSEA) and comparative fit index (CFI), with RMSEA values of ≤ 0.08 and CFI values ≥ 0.90 considered as indicating acceptable fit.[25] Details on the factor-of-curves building process can be found in Supplementary II.

To estimate the relationship between initial level (intercept) and change over time (slope) of pain and cognitive function, we fitted three parallel process LGCMs. All three models included: a correlation between the intercept of pain and the intercept of cognitive function; a regression path from intercept of pain to the slope of pain; and regression paths from the intercept of cognitive function, the intercept of pain, and the slope of pain to the slope of cognitive function. The structure of the regression paths is based on our hypothesis and temporality of the effects. We did not want to assume a directional relationship between baseline pain severity and baseline cognitive function because both directions have been reported and correlational associations have previously been suggested in the literature.[15,26] Model 1 was unadjusted, model 2 adjusted for age and sex, and model 3 additionally adjusted for socioeconomic status, ethnicity and comorbidities. Covariates were all considered time invariant and added using regression paths from each covariate to the intercepts and slopes of both pain and cognitive function. A simplified path diagram of the parallel process model is shown in Figure 1. Here, we are interested in how pain relates to cognitive decline. Our primary focus is the regression path from the slope of pain to slope of cognitive function, and the secondary focus is the regression path from intercept of pain to slope of cognitive function (both paths shown in red in Figure 1).

Since we modelled these regression paths simultaneously, the resultant path estimates for the intercept and the slope of pain on cognitive declines are mutually conditioned. That is, the contributions of pain at baseline on cognitive change is independent of the statistical contribution of changes in pain, and vice versa. Thus, where both paths are significant, this indicates unique, independent contributions of both how much pain one has to begin with, and how much that changes over time, to cognitive ageing trajectories. We also ran separate supplementary models where the intercept and slope of pain are not mutually conditioned on each other to highlight the additive process of our model design.

Missing data is common in longitudinal studies, often resulting from participants not answering questions, intermittently skipping waves or dropping out of the study completely. ELSA refreshes its sample by adding new participants at certain waves to maintain its representativeness of the English population aged over 50 years. Much of the missing data in our analysis arose by design: refresher participants had missing data on the previous waves, the animal naming test was not administered at wave 6 and the letter cancelling test was only assessed up to wave 5. Patterns of missingness and rate of attrition for the pain variable are reported in Supplementary III. In this study, we used the maximum likelihood estimator with robust standard errors (MLR), specifying that the pain severity variables were ordered categorical. The *probit* link function was used, which is appropriate for categorical data with more than two levels. With this estimator, our model ran under Full Information Maximum Likelihood (FIML) which uses all available data points to estimate model parameters. FIML assumes our data is missing at random (MAR);[46,50] that is, after accounting for the observed variables in the model, the probability that a value is missing does not depend on the unobserved values. This assumption allows FIML to produce unbiased parameter estimates by utilising all available data points.

If there were missing data on the covariates, our software removed the participants from the study. This led to a small discrepancy in sample size in our three models (model 1 n = 18,880, model 2 n = 18,861, model 3 n = 18,462).

To achieve convergence of the categorical LGCM of pain severity under the MLR estimator and *probit* link function and due to low representation in the *severe* category of pain severity (cross-wave average N = 685 (8%)), we applied partial invariance of the thresholds: the third threshold was free to vary over time whilst the first and second thresholds were held invariant. It is not possible to compute conventional fit indices such as RMSEA and CFI after including the latent factors derived from the categorical pain variable. However, inspection of the cognitive test loadings on the higher-order factors confirmed they remained similar to the factor-of-curves LGCM of general cognitive function before its integration into the parallel process model (see Supplementary IV for detailed table).

We also investigated the associations between the intercepts and slopes of pain and each individual cognitive test, following the same pattern of correlations and regression paths as the primary analysis but without the higher-order general cognitive function factors (illustrative path diagram shown in Supplementary V).

We used R version 4.5.1[39] in the RStudio environment version 2025.09.1+401[38] for data preparation and plotting. MPlus version 8.11[32] was used for SEM analysis. The R package MPlusAutomation version 1.2[17] allowed us to transfer factor scores from MPlus to R. See Acknowledgements for code repository details.

## RESULTS

### Descriptive statistics

Detailed descriptive results of the pain and cognitive function variables at each wave are shown in Table 1 (plots showing their distribution are in Figure 2A and Supplementary I). The combined sample of 19,376 participants had a mean age of 58.84 years at wave 1 and 53% were female. Other participant characteristics are reported in Table 2 and more details on wave-by-wave characteristics are available in Supplementary VI.

**Table 1:** Descriptive statistics on the three cognitive tests and reports of pain severity. *Time since wave 1 is measured in years and corresponds to the first order factor loadings.

| Wave number | 1 | 2 | 3 | 4 | 5 | 6 | 7 | 8 | 9 | 10 | 11 |
| --- | --- | --- | --- | --- | --- | --- | --- | --- | --- | --- | --- |
| Time since wave 1* | 0 | 2.29 | 4.29 | 6.25 | 8.29 | 10.21 | 12.21 | 14.21 | 16.29 | 19.79 | 21.59 |
| N | 12039 | 9380 | 9662 | 10928 | 10153 | 10372 | 9195 | 8060 | 7286 | 6010 | 5690 |
| **Recall** |  |  |  |  |  |  |  |  |  |  |  |
| N | 11645 | 9187 | 9359 | 10416 | 9562 | 9680 | 8432 | 7425 | 6635 | 5307 | 5018 |
| Mean | 9.46 | 9.94 | 10.29 | 10.39 | 10.41 | 10.65 | 10.46 | 10.57 | 10.47 | 10.40 | 10.62 |
| SD | 3.59 | 3.62 | 3.70 | 3.65 | 3.73 | 3.72 | 3.79 | 3.79 | 3.71 | 3.54 | 3.53 |
| **Animal Naming** |  |  |  |  |  |  |  |  |  |  |  |
| N | 11618 | 9150 | 9312 | 10366 | 9461 |  | 8322 | 7382 | 6598 | 5339 | 5071 |
| Mean | 19.38 | 19.90 | 20.31 | 20.78 | 20.94 |  | 21.29 | 21.59 | 22.01 | 22.18 | 22.37 |
| SD | 6.35 | 6.47 | 6.78 | 6.76 | 6.69 |  | 6.87 | 6.99 | 7.17 | 7.29 | 7.28 |
| **Letter cancellation** |  |  |  |  |  |  |  |  |  |  |  |
| N | 11237 | 8913 | 8897 | 9341 | 8935 |  |  |  |  |  |  |
| Mean | 18.74 | 18.44 | 18.94 | 19.10 | 18.89 |  |  |  |  |  |  |
| SD | 5.96 | 5.85 | 5.74 | 5.47 | 5.43 |  |  |  |  |  |  |
| **Pain severity** |  |  |  |  |  |  |  |  |  |  |  |
| None |  |  |  |  |  |  |  |  |  |  |  |
| N | 7343 | 5749 | 5843 | 6315 | 5766 | 5608 | 4986 | 4349 | 3860 | 3034 | 2850 |
| % | 62.0 | 62.1 | 62.1 | 60.4 | 60.1 | 57.9 | 59.0 | 58.1 | 57.8 | 55.5 | 55.0 |
| Mild |  |  |  |  |  |  |  |  |  |  |  |
| N | 1252 | 972 | 1085 | 1242 | 1168 | 1227 | 1069 | 949 | 876 | 786 | 718 |
| % | 10.6 | 10.5 | 11.5 | 11.9 | 12.2 | 12.7 | 12.6 | 12.7 | 13.1 | 14.4 | 13.9 |
| Moderate |  |  |  |  |  |  |  |  |  |  |  |
| N | 2177 | 1817 | 1725 | 2090 | 1873 | 2084 | 1771 | 1545 | 1451 | 1204 | 1202 |
| % | 18.4 | 19.6 | 18.3 | 20.0 | 19.5 | 21.5 | 20.9 | 20.7 | 21.7 | 22.0 | 23.2 |
| Severe |  |  |  |  |  |  |  |  |  |  |  |
| N | 1077 | 714 | 754 | 816 | 792 | 770 | 628 | 637 | 493 | 445 | 411 |
| % | 9.1 | 7.7 | 8.0 | 7.8 | 8.3 | 7.9 | 7.4 | 8.5 | 7.4 | 8.1 | 7.9 |

**Table 2:** Covariates descriptive characteristics. SES = Socioeconomic Status based on the National Statistics Socio-Economic Classification.[25] Comorbidities are considered if participant ever reported being told by a clinician that they have the diagnosis at any wave. Values are mean (SD) for age and N (%) for all other variables.

|  | Mean (SD) / N (%) | Missing N (%) |
| --- | --- | --- |
| Age | 58.84 (11.9) | 7 (<0.1%) |
| Sex |  | 20 (0.1%) |
| Female | 10,355 (53.4%) |  |
| Male | 9,021 (46.6%) |  |
| Ethnicity |  | 114 (0.6%) |
| White | 18,420 (95.1%) |  |
| Non-white | 956 (5.9%) |  |
| Socioeconomic status |  | 672 (3.5%) |
| SES 1 | 1,933 (10.0%) |  |
| SES 2 | 3,892 (20.1%) |  |
| SES 3 | 2,893 (14.9%) |  |
| SES 4 | 1,943 (10.0%) |  |
| SES 5 | 1,907 (9.8%) |  |
| SES 6 | 3,158 (16.3%) |  |
| SES 7 | 2,746 (14.2%) |  |
| SES 8 | 232 (1.2%) |  |
| Comorbidities |  |  |
| Diabetes | 1,121 (5.8%) | 13 (<0.1%) |
| Hypertension | 3,444 (17.8%) | 13 (<0.1%) |
| Heart disease | 2,402 (12.4%) | 15 (<0.1%) |
| Lung disease | 839 (4.3%) | 14 (<0.1%) |
| Stroke | 712 (3.7%) | 13 (<0.1%) |
| Cancer | 1,285 (6.6%) | 13 (<0.1%) |

**Figure 2:**
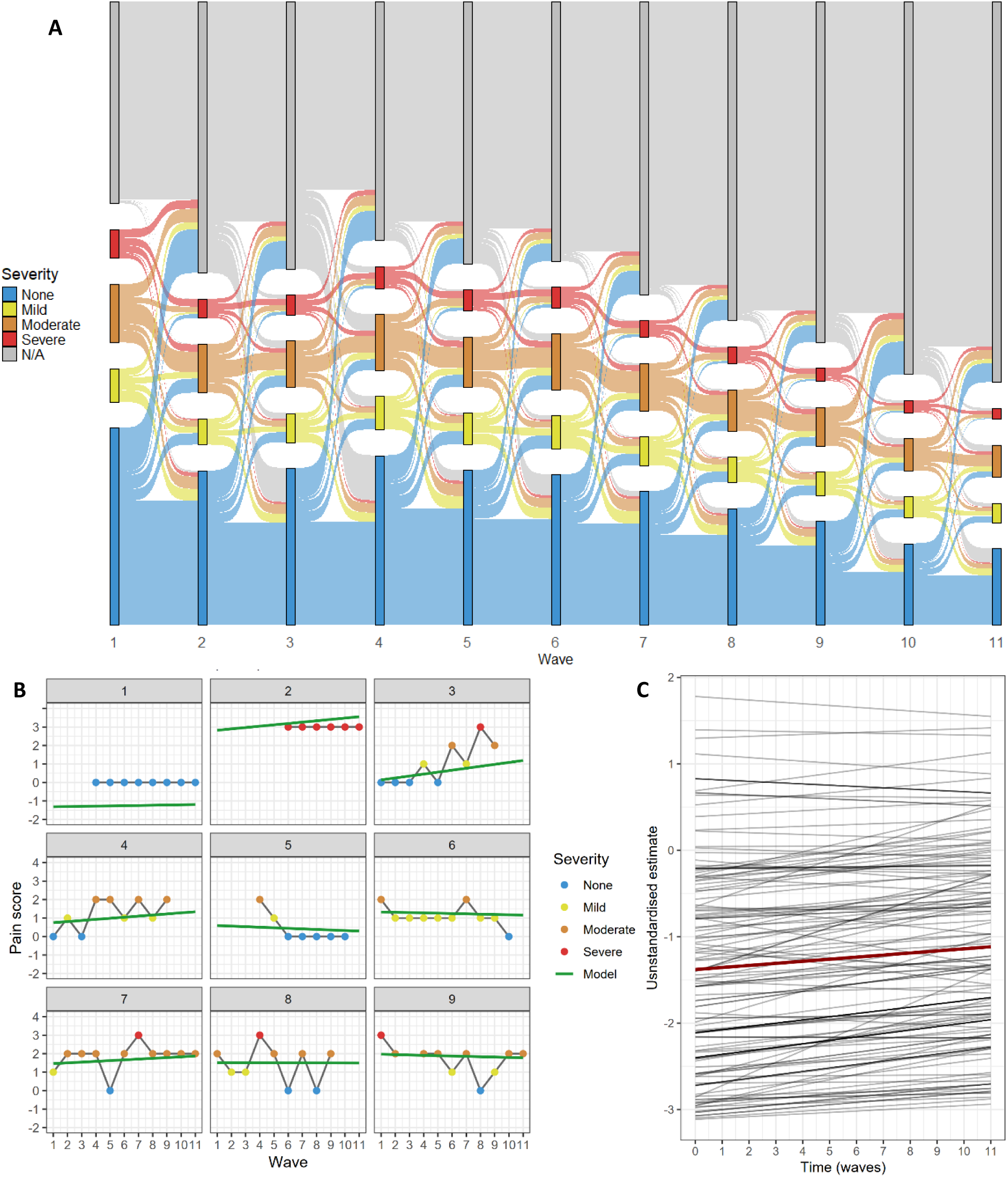
Trajectories of pain severity. A: Sankey diagram of pain severity with category change over time in the entire sample. B: Nine participants’ pain reports over 11 waves and their model-estimated trajectories. These nine participants were selected for illustrative purposes and are not systematically representative of the study sample. The green lines labelled “Model” are the individual-level model-estimated trajectories. C: The unstandardised estimates of pain severity intercept and slope from a random sample of 150 participants estimated by the measurement model are shown in grey, and the mean unstandardised estimate is shown in dark red (intercept β = −1.381, slope β = 0.024).

### Measurement models

#### Pain severity univariate LGCM

There was substantial variation in the reported level and change in pain over time as shown in the Sankey diagram in Figure 2A. Some participants reported no pain at all throughout, some reported severe pain at all waves, some showed decreasing or increasing levels of pain across time, and, in participants with pain, severity could fluctuate from one wave to the next. Figure 2B shows an illustrative example of 9 participants’ pain reports over 11 waves, chosen to demonstrate the diversity of pain trajectories, along with the model estimated trajectories.

The LGCM of pain severity does not provide standard fit indices due to the MLR estimator having to compute latent factors from categorical variables. The standardised estimates demonstrate that, on average, the participants report worsening pain over time. Although the third threshold was free to vary across waves, it remained within a limited range of values (min = 1.029, max = 1.409). Figure 2C shows the model-estimated trajectories of pain severity, with the average trajectory depicted in red and 150 randomly selected individual-level trajectories depicted in grey. This underscores the high variance in the initial pain severity and lower variance in the slope of pain over time.

#### Cognitive function univariate LGCM

The individual LCGM of cognitive tests all demonstrated excellent fit (Word recall CFI = 0.956, RMSEA = 0.039; Animal naming CFI = 0.976, RMSEA = 0.026; Letter cancellation CFI = 0.989, RMSEA = 0.017) and, on average, a decline in test performance over time (Word recall intercept β = 3.485, slope β = −.5 6; Animal naming intercept β = 3.8 6, slope β = −.45; Letter cancellation intercept β = 4. 78, slope β = −0.316; all estimates significant with *p* ≤ 0.008). Model-estimated average and individual-level trajectories for 150 randomly selected participants for each test are shown in Supplementary VII.

The factor-of-curves model of cognitive function showed excellent fit (CFI = 0.973, RMSEA = 0.021) and estimated an average decrease over time in general cognitive performance (standardised estimates for intercept β = 3.978, p < 0.001 and slope β = −0.754, p < 0.001). Model-estimated average and individual-level trajectories for 150 randomly selected participants are shown in Figure 3. Given that the three cognitive tests capture different domains of cognitive function (memory, executive function and processing speed), variation in the magnitude of the estimated loadings is expected and indicate a different degree of contribution to the higher-order factor. The general cognitive function factors captured nearly all the variance in both word recall (standardised loading on intercept λ = 0.896 and slope λ = 0.853) and animal naming (standardised loading on intercept λ = 0.828 and slope λ = .98). In contrast, the letter cancellation’s loadings contributed moderately to both higher-order factors (λ = 0.665 on the intercept and λ = 0.633 on the slope, in standardised units), meaning that a greater proportion of its variance is not shared with the other two tests. This could also be a result of this variable’s high amount of missing data by design or it might reflect on the quality of the test.

**Figure 3:**
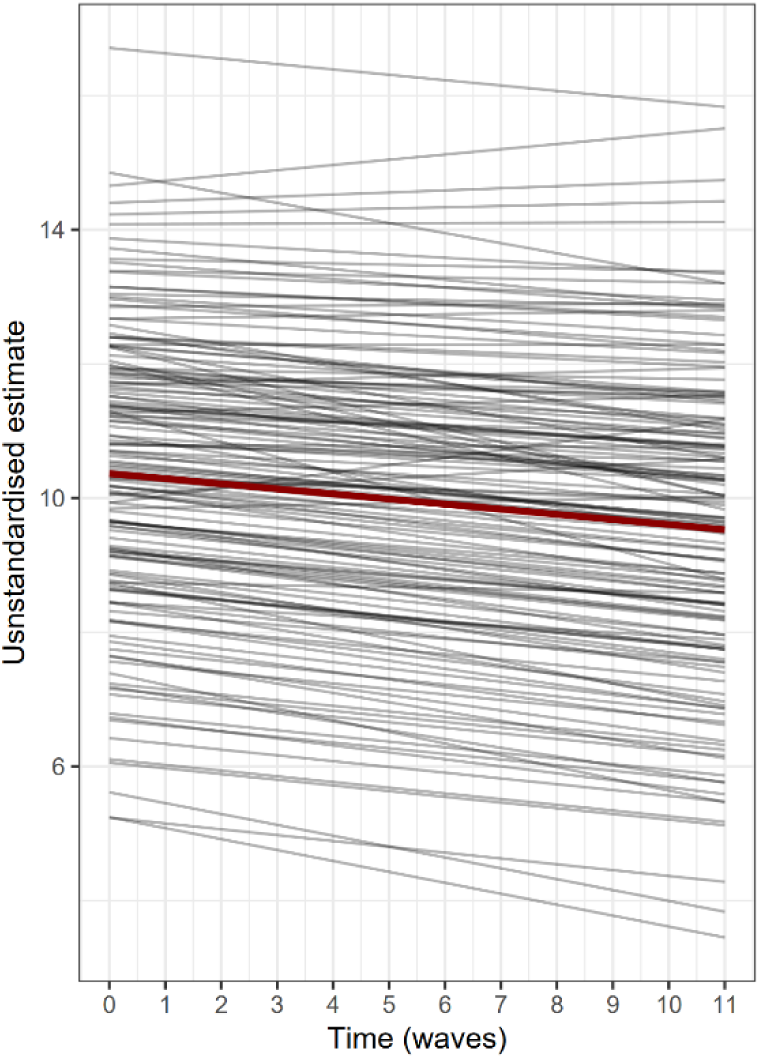
Trajectories of general cognitive function. The unstandardised estimates of general cognitive function intercept and slope from a random sample of 150 participants estimated by measurement model are shown in grey, and the mean unstandardised estimate is shown in dark red (intercept β = 10.364, slope β = −0.076).

Evaluation of rank order stability of pain and cognitive function demonstrated high correlations across waves; indicating that participants’ baseline rank tends to be maintained across time (see Supplementary VIII).

We also conducted sensitivity analyses to investigate the robustness of our findings. When inspecting the means of the latent variables across models, the mean intercept and slope for cognitive function stayed relatively consistent; however, the means of the latent variables of pain severity showed somewhat greater variability. We therefore report them alongside each model result.

### Longitudinal associations between pain and cognitive function (parallel process LGCM)

We report below results in standardised units and p-values have undergone false discovery rate correction following the Benjamini–Hochberg procedure on each individual model. Summary results for the relationship between pain severity and cognitive function intercept and slope are shown in Figure 4 and Table 3. More detailed results including regression coefficients on the covariates are reported in Supplementary IX, model fit information in Supplementary X and longitudinal associations between the latent pain factors and individual test intercepts and slopes are described in Supplementary XI.

**Table 3:** Summary of results. Estimates are all standardised. β = Estimate, SE = standard error, *p* = p-value, g = general cognitive function as determined by the factor-of-curves method. → represent regression paths, ↔ show correlations. The p-values are adjusted within each model for false discovery rate using the Benjamini–Hochberg procedure with α = 0.05.

| Variables | Model 1 (no covariates) |  |  | Model 2 (age and sex) |  |  | Model 3 (age, sex, ethnicity, socio-economic status and comorbidities) |  |  |
| --- | --- | --- | --- | --- | --- | --- | --- | --- | --- |
| | $\beta$ | SE | $p$ | $\beta$ | SE | $p$ | $\beta$ | SE | $p$ |
| **General cognitive function** |  |  |  |  |  |  |  |  |  |
| Slope of pain → Slope of g | -0.096 | 0.030 | 0.001 | -0.053 | 0.026 | 0.039 | -0.025 | 0.026 | 0.365 |
| Intercept of pain → Slope of g | -0.126 | 0.020 | <0.001 | -0.117 | 0.018 | <0.001 | -0.104 | 0.018 | <0.001 |
| Intercept of g → Slope of g | 0.018 | 0.035 | 0.604 | -0.145 | 0.031 | <0.001 | -0.206 | 0.036 | <0.001 |
| Intercept of pain → Slope of pain | -0.410 | 0.023 | <0.001 | -0.431 | 0.023 | <0.001 | -0.461 | 0.023 | <0.001 |
| Intercept of g ↔ Intercept of pain | -0.259 | 0.011 | <0.001 | -0.240 | 0.012 | <0.001 | -0.156 | 0.013 | <0.001 |
| Mean intercept of g | 3.978 | 0.045 | <0.001 | 3.821 | 0.043 | <0.001 | 3.715 | 0.062 | <0.001 |
| Mean slope of g | -0.748 | 0.200 | <0.001 | -0.426 | 0.134 | 0.002 | -0.246 | 0.210 | 0.312 |
| Mean intercept of pain | 0.176 | 0.033 | <0.001 | -0.506 | 0.034 | <0.001 | -0.015 | 0.069 | 0.823 |
| Mean slope of pain | 0.517 | 0.027 | <0.001 | 0.173 | 0.033 | <0.001 | 0.373 | 0.089 | <0.001 |

**Figure 4:**
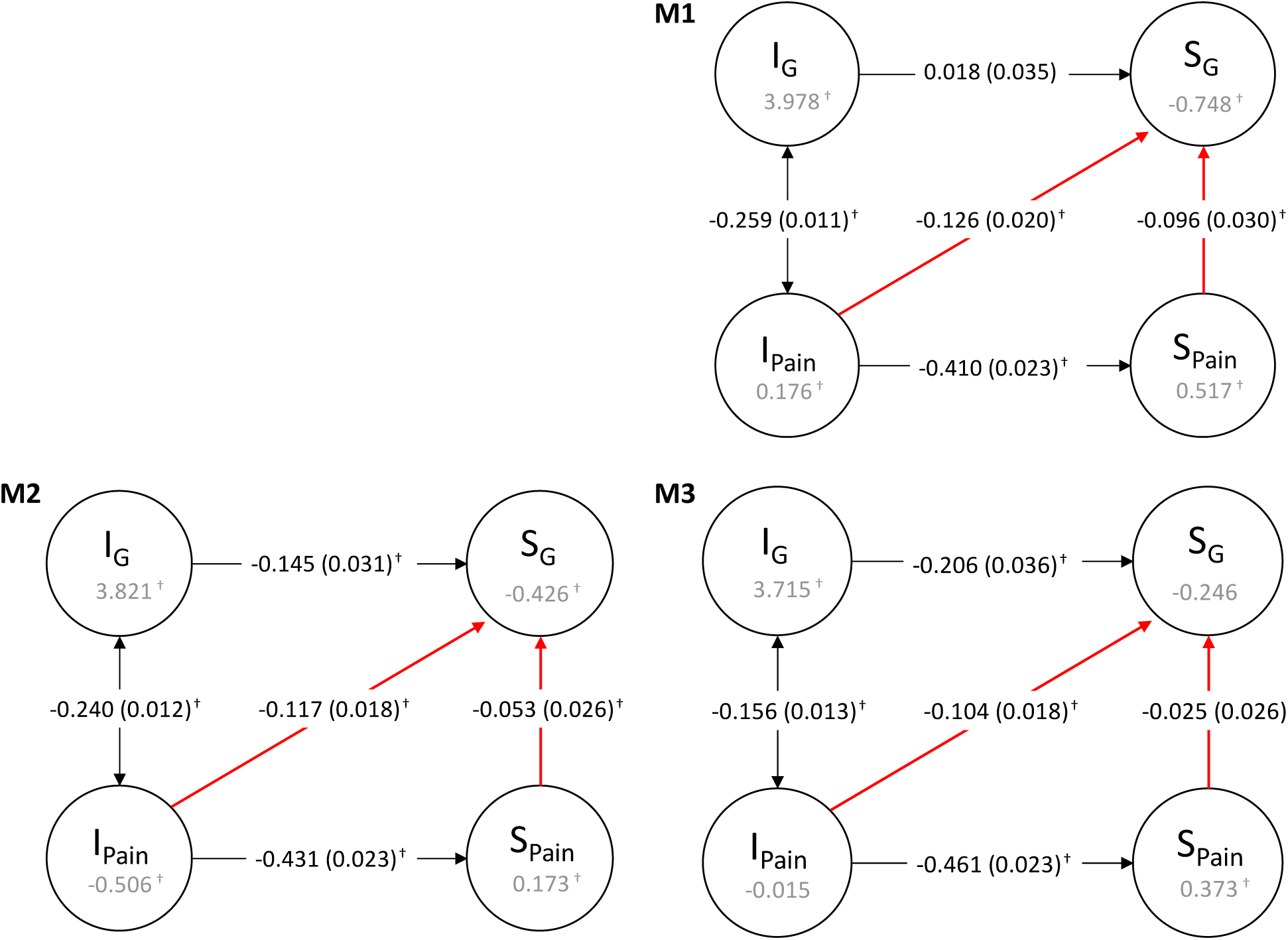
Simplified path diagram of parallel process LGCM of pain severity and cognitive function. Model 1 (M1) has not been adjusted for any covariates, model 2 (M2) was adjusted for age and sex, and model 3 (M3) was adjusted for age, sex, ethnicity, socioeconomic status and comorbidities. I = Intercept, S = Slope, G = General cognitive function. The values inside the circles are the mean estimates. Single-headed arrows are regression paths and double-headed arrows are correlation paths. All estimates are standardised. Standard errors are in brackets and † indicates a statistically significant value at α = 0.05 after false discovery rate correction following the Benjamini– Hochberg procedure.

In the unadjusted model (M1), more initial pain was correlated with lower initial cognitive function (*r* = −0.259, *p* < .1) and with accelerated cognitive decline (β = −0.126, *p* < 0.001). More initial pain was strongly associated with worsening pain over time (β = −0.410, *p* < 0.001). Increasing pain severity over time was associated with a faster rate of cognitive decline over time (β = −0.096, *p* = 0.001). Since the model is simultaneously accounting for the effect of initial pain on slopes of pain and *g*; worsening pain over time has an effect on the rate of cognitive decline above and beyond the effect of initial pain. When adjusting for age and sex (M2), pain severity slope remained associated with the slope of cognitive function (β = −0.053, *p* = 0.039), higher levels of initial pain remained correlated with lower initial cognitive function (*r* = −0.240, *p* < 0.001), and remained associated with worsening pain over time (β = −0.431, *p* < .1) and accelerated cognitive decline (β = −0.117, *p* < 0.001). When additionally adjusting for ethnicity, socioeconomic status, and comorbidities (M3), the magnitude of the association between pain slope and general cognitive function slope was attenuated to non-significance (β = −0.025, *p* = 0.365). In this fully-adjusted model, higher initial pain severity remained significantly correlated with lower initial cognitive function (*r* = 0.156, p < 0.001) and remained associated with worsening pain over time (β = −0.461, *p* < 0.001) and with steeper cognitive decline over time (β = −0.104, p < 0.001).

### Sensitivity analyses

To explore our results further, we ran two bivariate models where the intercept and slope of pain were not mutually conditioned. We found that the path estimates of these factors on cognitive change remained significant and of similar magnitude (see Supplementary XII)

To identify which covariates may be driving the attenuation of the slope-slope association in model 3, we fitted nested models adding each set of covariates (i.e. comorbidities, socioeconomic status and ethnicity) individually to the age- and sex-adjusted model (M2): The addition of ethnicity to model 2 did not affect the estimate (β = −0.054, p = 0.036); in contrast, both socioeconomic status and comorbidities significantly attenuated the regression of slope of cognitive function on slope of pain (β = −.35, p = .17 and β = −0.042, p = 0.107 respectively). In all three of these nested models, the standard error of the slope-slope coefficient remained constant (SE = 0.026), indicating no loss of power or precision.

We also tested for non-linear trajectories for cognitive function and found that a linear model seemed to fit the data well and avoided overfitting. Adding a quadratic term to the LGCMs of individual cognitive tests showed relatively similar fit on the three tests (word recall linear: CFI = 0.956, RMSEA 0.039, quadratic: CFI 0.983, RMSEA 0.025 – animal naming linear: CFI 0.976, RMSEA 0.026, quadratic: CFI 0.990, RMSEA 0.018 - letter cancellation linear: CFI 0.991, RMSEA 0.020, quadratic: CFI 0.991, RMSEA 0.019) however, MPlus was unable to estimate standard errors when combined into a quadratic factor-of-curves.

To assess the robustness of the results, we ran two more sets of parallel process LGCM models: the first was run without partial invariance of the thresholds for the pain severity categorical variable; and for the second set of models, we used only waves 1 to 9 to examine the effect of pain severity on cognitive function before the COVID-19 pandemic (the time elapsed between waves 9, 10 and 11 became more irregular). Both sets of models showed very similar regression and correlation path coefficients to the primary analysis (see Supplementary XIII).

## DISCUSSION

In this study of nearly 20,000 middle-aged and older adults residing in England, followed for more than 20 years, we examined how longitudinal variations in pain severity relate to changes in cognitive function over time. We found evidence that worsening pain over time was associated with steeper cognitive decline, independently of baseline pain level, and partially attenuated by age and sex. This association was further attenuated and non-significant when additionally adjusting for socioeconomic status (SES), ethnicity and comorbidities although downstream analyses indicate that ethnicity did not contribute to the attenuation. Our models also indicate that initial levels of pain severity were consistently associated with faster rates of decline in cognitive function, even when adjusting for sociodemographic and health variables. We conclude that both the intensity of initial pain and its worsening over time relate to a steeper decline in cognition independently of each other; however, the association between increasing pain severity and cognitive decline is partly explained by the cumulative effects of pain at baseline, sex, age, SES and comorbid chronic conditions.

The present work is one of only a few studies to have examined change in both chronic pain and cognitive function over time. Our findings substantiate the conclusions drawn by He et al.[20] in their study of ELSA (n = 5,685) and the Health and Retirement Study (n = 7,619). Using group-based trajectory modelling and linear mixed effects, they found that the high-stable and moderate-increasing categories of probability of pain presence estimated over 8-10 years were associated with steeper cognitive decline over the subsequent 8-12 years. We expand these results by capturing variations in pain severity rather than pain presence, and by utilising LGCM to directly link within-person change in pain to within-person change in cognitive function over a 22-year time period. Within-person changes were also captured by Milani et al.[30] in their cross-lagged analysis of older Puerto Rican adults. They too found greater cognitive decline in participants who had pain at follow-up compared to those who did not; however, this association was bidirectional and only measured over two waves of data. Furthermore, over one third of their sample participants were excluded from the analysis due to missing data whereas our study was able to retain its large sample size by leveraging SEM’s Full Information Maximum Likelihood (FIML) method. To the best of our knowledge, ours is the first study to examine the association of pain and cognitive function using LGCM and FIML, thereby maintaining representativeness and reducing selection bias.

Another key strength ensuing from our use of LGCM is the factor-of-curves which has several methodological advantages over previous studies. Techniques which average standardised scores – such as He et al.’s – to create a global measure of cognitive function implicitly treat each test as equally reliable, fail to account for measurement error, and discount the shared structure of cognitive function. In contrast, the factor-of-curves method allows us to simultaneously estimate latent intercepts and slopes from multiple cognitive tests and aggregate these into a latent higher-order cognitive factor. This approach not only explicitly accounts for measurement error in repeated indicators but also accurately captures the well-replicated hierarchical structure of cognitive changes over time.[2,18,42,56] Thus, our method provides a more robust, theoretically sound, and psychometrically valid estimation of cognitive change than was previously achieved.

As one of the largest studies with one of the longest follow-ups in the literature, our work also consolidates the evidence on the predictive nature of cross-sectional measures of pain on cognitive decline. [6,22,23,45,58][43] In many of these studies, pain is defined as chronic or persistent by design, either measuring it twice or recording pain which had lasted at least 3 months. On the other hand, Van der Leeuw et al.[28] investigated pain over a shorter period of time and found no association with cognitive decline; therefore our measure of baseline pain severity is likely capturing a chronic characteristic of the condition. This stable, long-term history of pain could be the primary pan-related influence on cognitive decline, potentially through mechanisms such as neurostructural changes[36] and inflammation. This was observed by Sadlon et al.[47] who found an increase in inflammatory markers in the blood and cerebrospinal fluid of patients reporting pain. In contrast, within-person changes in pain over time were modest, as is typically observed in longitudinal studies, and small relative to baseline variance. Consequently, any effects on cognitive decline produced by change in pain, if present, are likely to be subtle and potentially difficult to detect.

The use of the ELSA dataset for this study is another of its strength. Beyond its large sample size, ensuring sufficient statistical power to find associations even in complex modelling, the ELSA cohort is nationally representative of adults aged over 50 years living in England and continued follow up in participants who moved from the community to care homes. The capture of pain and cognitive function in a small number of care home residents mean our results are generalisable to frailer groups of older adults among the English population. Another strength of the ELSA study is its measure of pain. “Are you often troubled by pain?” does not specify a time frame or require participants to answer if they are currently experiencing pain. In their work on disability, Banks et al.[4] found that although 60% of adult Dutch respondents reported pain in the last thirty days, only a quarter of the sample answered yes to “are you often troubled with pain”. This phrasing might therefore selectively omit transient or minor episodes and more reliably capture persistent pain. Conversely, the use of a 4-point scale to measure pain severity might also be a limitation. This relatively crude measure may be less sensitive to subtle but important fluctuations in pain over time in contrast with the commonly used 11-point (0-10) numerical rating scale.[9,34,37] However, the low counts in the severe pain category within ELSA led the estimator to struggle with the transformation from categorical observed variable to continuous latent factors and compelled us to allow partial invariance of the pain thresholds. This likely drove the variability we observed in mean estimates of the pain intercept and slope across the several models run in sensitivity analysis. Nonetheless, estimated correlation and regression paths from pain intercept and slope to cognitive function intercept and slope were remarkably consistent across all parallel process models conducted; further affirming the conclusions drawn in the present study.

As one of the first studies to fully explore the longitudinal relationship between pain severity and cognitive decline, we selected covariates parsimoniously, however ELSA’s design was also restrictive. For example, we were unable to compute BMI: height and weight were scarcely measured across waves so a large number of participants would have been excluded from the study had we adjusted for it. To avoid selection bias in our conclusions, we decided instead to adjust for comorbidities which were likely to highly correlate with BMI such as diabetes, hypertension and heart disease. As per our downstream analyses, these comorbidities and the three others we included in this block of covariates (stroke, cancer and lung disease), seemed to weaken the association of worsening pain severity with steeper cognitive decline to non-significance. Socioeconomic status also had a considerable attenuating effect on this association. Mediation analyses were beyond the scope of this study; however, applied to these two variables, they may point to a potential mechanism underlying the conclusions of our analyses. Future research should investigate potential pathways through which pain is associated with cognitive decline, including other variables such as sleep disturbances, psychiatric conditions and prescription medications.

This study focused on only one measure of worsening pain over time; namely, increasing pain severity. Other indicators of worsening pain, such as duration of pain, the spread of pain to additional body sites, or increasing pain interference, may demonstrate a different pattern of association with cognitive decline. Pain interference (i.e., whether pain interferes with the ability to carry out work or household chores) is a strong predictor of cognitive impairment[5,24] but was not collected in the ELSA cohort. Investigating these other measures of pain over time and their association with cognitive function in similar fashion to the present study would be an important next step in understanding the longitudinal relationship between pain and cognitive function.

## CONCLUSION

Persistent pain can be debilitating and is associated with many negative health outcomes. In a sample of nearly 20,000 middle-aged and older adults, followed up for over 20 years, we found that increasing pain over time was associated with steeper rates of cognitive decline, and partly attenuated by sociodemographic variables and concurrent health conditions. Our study shows that higher initial pain is associated with accelerated cognitive decline, further elucidating the cognitive consequences of chronic pain and highlighting the importance of preventing its onset.

## Data Availability

Data from the ELSA dataset is available upon request from the UK Data Service. MPlus code used in the development of the SEM models is available at https://doi.org/10.5281/zenodo.18593947 under the Creative Commons Attribution 0.4 International License (CC-BY-4.0).

## ACKNOWLEDGEMENTS

The authors have no conflicts of interest to declare.

This research was funded by the Legal & General Group (research grant to establish the independent Advanced Care Research Centre at University of Edinburgh). The funder had no role in conduct of the study, interpretation, or the decision to submit for publication. The views expressed are those of the authors and not necessarily those of Legal & General.

SRC was supported by a Sir Henry Dale Fellowship, jointly funded by the Wellcome Trust and the Royal Society (221890/Z/20/Z), and also by the US National Institutes of Health (National Institute on Aging; 1RF1AG073593).

Research reported in this publication was supported by the National Institute On Aging of the National Institutes of Health under Award Number R01AG017644. The content is solely the responsibility of the authors and does not necessarily represent the official views of the National Institutes of Health. ELSA is funded by the NIHR Policy Research Programme (HEI) 198_1074_03. The views expressed are those of the author(s) and not necessarily those of the NIHR or the Department of Health and Social Care.[3]

We are grateful to the participants of ELSA for their valuable contributions over the years which made this work possible.

No original datasets were generated over the course of this research. ELSA data is managed by the UK Data Service and access for researchers may be requested through their platform. MPlus code used in the development of the SEM models is available at https://doi.org/10.5281/zenodo.18593947 under the Creative Commons Attribution 0.4 International License (CC-BY-4.0).

## REREFENCES

[1] de Aguiar GPCG, Saraiva MD, Khazaal EJB, de Andrade DC, Jacob-Filho W, Suemoto CK. Persistent pain and cognitive decline in older adults: a systematic review and meta-analysis from longitudinal studies. PAIN 2020;161:2236.

[2] Altschul DM, Starr JM, Deary IJ. Cognitive function in early and later life is associated with blood glucose in older individuals: analysis of the Lothian Birth Cohort of 1936. Diabetologia 2018;61:1946–1955.

[3] Banks J, Batty GD, Breedvelt J, Coughlin K, Crawford R, Marmot M, Nazroo J, Oldfield Z, Steel N, Steptoe A, Wood M, Zaninotto P. English Longitudinal Study of Ageing: Waves 0-11, 1998-2024. 2025. doi:10.5255/UKDA-SN-5050-34.

[4] Banks J, Kapteyn A, Smith JP, Soest A van, Cutler DM, Wise DA eds. Work Disability is a Pain in the ****, Especially in England, the Netherlands, and the United States. Health at Older Ages: The Causes and Consequences of Declining Disability Among the Elderly. University of Chicago Press, 2009. p. 0. doi:10.7208/chicago/9780226132327.003.0010.

[5] Bell T, Franz CE, Kremen WS. Persistence of pain and cognitive impairment in older adults. J Am Geriatr Soc 2022;70:449–458.

[6] Bell TR, Sprague BN, Ross LA. Longitudinal associations of pain and cognitive decline in community-dwelling older adults. Psychol Aging 2022;37:715–730.

[7] Berryman C, Stanton TR, Bowering JK, Tabor A, McFarlane A, Moseley LG. Evidence for working memory deficits in chronic pain: A systematic review and meta-analysis. PAIN 2013;154:1181.

[8] Berryman C, Stanton TR, Bowering KJ, Tabor A, McFarlane A, Moseley GL. Do people with chronic pain have impaired executive function? A meta-analytical review. Clin Psychol Rev 2014;34:563– 579.

[9] Breivik H, Borchgrevink PC, Allen SM, Rosseland LA, Romundstad L, Hals EKB, Kvarstein G, Stubhaug A. Assessment of pain. Br J Anaesth 2008;101:17–24.

[10] Carroll JB ed. Higher-Order Factors of Cognitive Ability. Human Cognitive Abilities: A Survey of Factor-Analytic Studies. Cambridge: Cambridge University Press, 1993. pp. 577–628. doi:10.1017/CBO9780511571312.016.

[11] Corley J, Conte F, Harris SE, Taylor AM, Redmond P, Russ TC, Deary IJ, Cox SR. Predictors of longitudinal cognitive ageing from age 70 to 82 including APOE e4 status, early-life and lifestyle factors: the Lothian Birth Cohort 1936. Mol Psychiatry 2023;28:1256–1271.

[12] Dunn KM, Campbell P, Jordan KP. Long-term trajectories of back pain: cohort study with 7-year follow-up. BMJ Open 2013;3:e003838.

[13] Enthoven WTM, Koes BW, Bierma-Zeinstra SMA, Bueving HJ, Bohnen AM, Peul WC, van Tulder MW, Berger MY, Luijsterburg PAJ. Defining trajectories in older adults with back pain presenting in general practice. Age Ageing 2016;45:878–883.

[14] Fayaz A, Croft P, Langford RM, Donaldson LJ, Jones GT. Prevalence of chronic pain in the UK: a systematic review and meta-analysis of population studies. BMJ Open 2016;6:e010364.

[15] Ferreira K, Velly A. Cognitive Decline Overtime in Patients with Chronic Pain and Headache: a Systematic Review (P1-1.Virtual). Neurology 2022;98:651.

[16] Glette M, Stiles TC, Borchgrevink PC, Landmark T. The Natural Course of Chronic Pain in a General Population: Stability and Change in an Eight–Wave Longitudinal Study Over Four Years (the HUNT Pain Study). J Pain 2020;21:689–699.

[17] Hallquist MN, Wiley JF. MplusAutomation: An R Package for Facilitating Large-Scale Latent Variable Analyses in Mplus. Struct Equ Model Multidiscip J 2018;25:621–638.

[18] Hamilton OKL, Cox SR, Okely JA, Conte F, Ballerini L, Bastin ME, Corley J, Taylor AM, Page D, Gow AJ, Muñoz Maniega S, Redmond P, Valdés-Hernández M del C, Wardlaw JM, Deary IJ. Cerebral small vessel disease burden and longitudinal cognitive decline from age 73 to 82: the Lothian Birth Cohort 1936. Transl Psychiatry 2021;11:376.

[19] Hayden KM, Reed BR, Manly JJ, Tommet D, Pietrzak RH, Chelune GJ, Yang FM, Revell AJ, Bennett DA, Jones RN. Cognitive decline in the elderly: an analysis of population heterogeneity. Age Ageing 2011;40:684–689.

[20] He Z, Li G, Chen Z, Hu Z, Wang Q, Huang G, Luo Q. Trajectories of pain and their associations with long-term cognitive decline in older adults: evidence from two longitudinal cohorts. Age Ageing 2024;53:afae183.

[21] Higgins DM, Martin AM, Baker DG, Vasterling JJ, Risbrough V. The Relationship Between Chronic Pain and Neurocognitive Function: A Systematic Review. Clin J Pain 2018;34:262.

[22] Honda H, Ashizawa R, Kameyama Y, Yoshimoto Y. Chronic pain in older adults with disabilities is associated with cognitive impairment—a prospective cohort study. Psychogeriatrics 2025;25:e13210.

[23] Kazim MA, Strahl A, Moritz S, Arlt S, Niemeier A. Chronic pain in osteoarthritis of the hip is associated with selective cognitive impairment. Arch Orthop Trauma Surg 2023;143:2189–2197.

[24] Kelleher E, Hawley S, Delmestri A, Arden N, Tracey I, Soni A. P125 The relationship between mid-life pain and cognitive impairment: a prospective, population-based study. Rheumatology 2021;60:keab247.120.

[25] Kline RB. Principles and practice of structural equation modeling, 4th ed. New York, NY, US: The Guilford Press, 2016.

[26] Kumaradev S, Fayosse A, Dugravot A, Dumurgier J, Roux C, Kivimäki M, Singh-Manoux A, Sabia S. Timeline of pain before dementia diagnosis: a 27-year follow-up study. PAIN 2021;162:1578.

[27] Landmark T, Dale O, Romundstad P, Woodhouse A, Kaasa S, Borchgrevink P c. Development and course of chronic pain over 4 years in the general population: The HUNT pain study. Eur J Pain 2018;22:1606–1616.

[28] Liu X, Li L, Tang F, Wu S, Hu Y. Memory impairment in chronic pain patients and the related neuropsychological mechanisms: a review. Acta Neuropsychiatr 2014;26:195–201.

[29] McArdle JJ. Dynamic but Structural Equation Modeling of Repeated Measures Data. In: Nesselroade JR, Cattell RB, editors. Handbook of Multivariate Experimental Psychology. Boston, MA: Springer US, 1988. pp. 561–614. doi:10.1007/978-1-4613-0893-5_17.

[30] Milani SA, Bell TR, Crowe M, Pope CN, Downer B. Increasing Pain Interference Is Associated With Cognitive Decline Over Four Years Among Older Puerto Rican Adults. J Gerontol Ser A 2023;78:1005–1012.

[31] Moriarty O, McGuire BE, Finn DP. The effect of pain on cognitive function: A review of clinical and preclinical research. Prog Neurobiol 2011;93:385–404.

[32] Muthén LK, Muthén BO. Mplus User’s uide. 8th ed. Los Angeles, CA: Muthén & Muthén, 17 Available: https://www.statmodel.com/html_ug.shtml. Accessed 17 Oct 2025.

[33] NatCen Social Research, University College London. Health Survey for England. 2023. doi:10.5255/UKDA-SERIES-2000021.

[34] Nugent SM, Lovejoy TI, Shull S, Dobscha SK, Morasco BJ. Associations of Pain Numeric Rating Scale Scores Collected during Usual Care with Research Administered Patient Reported Pain Outcomes. Pain Med 2021;22:2235–2241.

[35] Office for National Statistics. The National Statistics Socio-economic classification (NS-SEC). n.d. Available: https://www.ons.gov.uk/methodology/classificationsandstandards/otherclassifications/thenationalstatisticssocioeconomicclassificationnssecrebasedonsoc2010. Accessed 16 Aug 2025.

[36] Pan P l., Zhong J g., Shang H f., Zhu Y l., Xiao P r., Dai Z y., Shi H c. Quantitative meta-analysis of grey matter anomalies in neuropathic pain. Eur J Pain 2015;19:1224–1231.

[37] Phillips S, Gift M, Gelot S, Duong M, Tapp H. Assessing the relationship between the level of pain control and patient satisfaction. J Pain Res 2013;6:683–689.

[38] Posit team. RStudio: Integrated Development Environment for R. 2024. Available: http://www.posit.co/.

[39] R Core Team. R: A Language and Environment for Statistical Computing. 2018. Available: https://www.R-project.org/.

[40] Rast P, Hofer SM. Longitudinal design considerations to optimize power to detect variances and covariances among rates of change: Simulation results based on actual longitudinal studies. Psychol Methods 2014;19:133–154.

[41] Raz N, Lindenberger U. Only time will tell: Cross-sectional studies offer no solution to the age– brain–cognition triangle: Comment on Salthouse (2011). Psychol Bull 2011;137:790–795.

[42] Ritchie SJ, Tucker-Drob EM, Cox SR, Corley J, Dykiert D, Redmond P, Pattie A, Taylor AM, Sibbett R, Starr JM, Deary IJ. Predictors of ageing-related decline across multiple cognitive functions. Intelligence 2016;59:115–126.

[43] Rong W, Zhang C, Zheng F, Xiao S, Yang Z, Xie W. Persistent moderate to severe pain and long-term cognitive decline. Eur J Pain 2021;25:2065–2074.

[44] Rouch I, Edjolo A, Laurent B, Pongan E, Dartigues J-F, Amieva H. Association between chronic pain and long-term cognitive decline in a population-based cohort of elderly participants. PAIN 2021;162:552.

[45] Rouch I, Edjolo A, Laurent B, Pongan E, Dartigues J-F, Amieva H. Association between chronic pain and long-term cognitive decline in a population-based cohort of elderly participants. PAIN 2021;162:552.

[46] Rubin DB. Inference and missing data. Biometrika 1976;63:581–592.

[47] Sadlon A, Takousis P, Ankli B, Alexopoulos P, Perneczky R, Initiative for the ADN. Association of Chronic Pain with Biomarkers of Neurodegeneration, Microglial Activation, and Inflammation in Cerebrospinal Fluid and Impaired Cognitive Function. Ann Neurol 2024;95:195–206.

[48] Salthouse TA. Selective review of cognitive aging. J Int Neuropsychol Soc 2010;16:754–760.

[49] Satorra A, Bentler PM. Ensuring Positiveness of the Scaled Difference Chi-square Test Statistic. Psychometrika 2010;75:243–248.

[50] Schafer JL, Graham JW. Missing data: Our view of the state of the art. Psychol Methods 2002;7:147–177.

[51] Schaie KW. What Can We Learn From Longitudinal Studies of Adult Development? Res Hum Dev 2005;2:133–158.

[52] Steptoe A, Breeze E, Banks J, Nazroo J. Cohort Profile: The English Longitudinal Study of Ageing. Int J Epidemiol 2013;42:1640–1648.

[53] Tucker-Drob EM. Global and domain-specific changes in cognition throughout adulthood. Dev Psychol 2011;47:331–343.

[54] Tucker-Drob EM. Global and domain-specific changes in cognition throughout adulthood. Dev Psychol 2011;47:331–343.

[55] Tucker-Drob EM, Brandmaier AM, Lindenberger U. Coupled cognitive changes in adulthood: A meta-analysis. Psychol Bull 2019;145:273–301.

[56] Tucker-Drob EM, Brandmaier AM, Lindenberger U. Coupled cognitive changes in adulthood: A meta-analysis. Psychol Bull 2019;145:273–301.

[57] Whitlock EL, Diaz-Ramirez LG, Glymour MM, Boscardin WJ, Covinsky KE, Smith AK. Association Between Persistent Pain and Memory Decline and Dementia in a Longitudinal Cohort of Elders. JAMA Intern Med 2017;177:1146–1153.

[58] Whitlock EL, Diaz-Ramirez LG, Glymour MM, Boscardin WJ, Covinsky KE, Smith AK. Association Between Persistent Pain and Memory Decline and Dementia in a Longitudinal Cohort of Elders. JAMA Intern Med 2017;177:1146–1153.

[59] Wickrama K, Lee TK, O’Neal CW, Lorenz F. Higher-Order Growth Curves and Mixture Modeling with Mplus: A Practical Guide. 2nd ed. New York: Routledge, 2021.

[60] Wilson RS, Beckett LA, Barnes LL, Schneider JA, Bach J, Evans DA, Bennett DA. Individual differences in rates of change in cognitive abilities of older persons. Psychol Aging 2002;17:179– 193.

[61] Zaninotto P, Steptoe A. English Longitudinal Study of Ageing. Encyclopedia of Gerontology and Population Aging. Springer, Cham, 2019. pp. 1–7. doi:10.1007/978-3-319-69892-2_335-1.

[62] Zhang X, Gao R, Zhang C, Chen H, Wang R, Zhao Q, Zhu T, Chen C. Evidence for Cognitive Decline in Chronic Pain: A Systematic Review and Meta-Analysis. Front Neurosci 2021;15. doi:10.3389/fnins.2021.737874.

