## Supplementary material for "Trajectories of pain and cognitive function: 22 years of evidence in mid-to-later life"

#### I. Distribution of the first-order variables

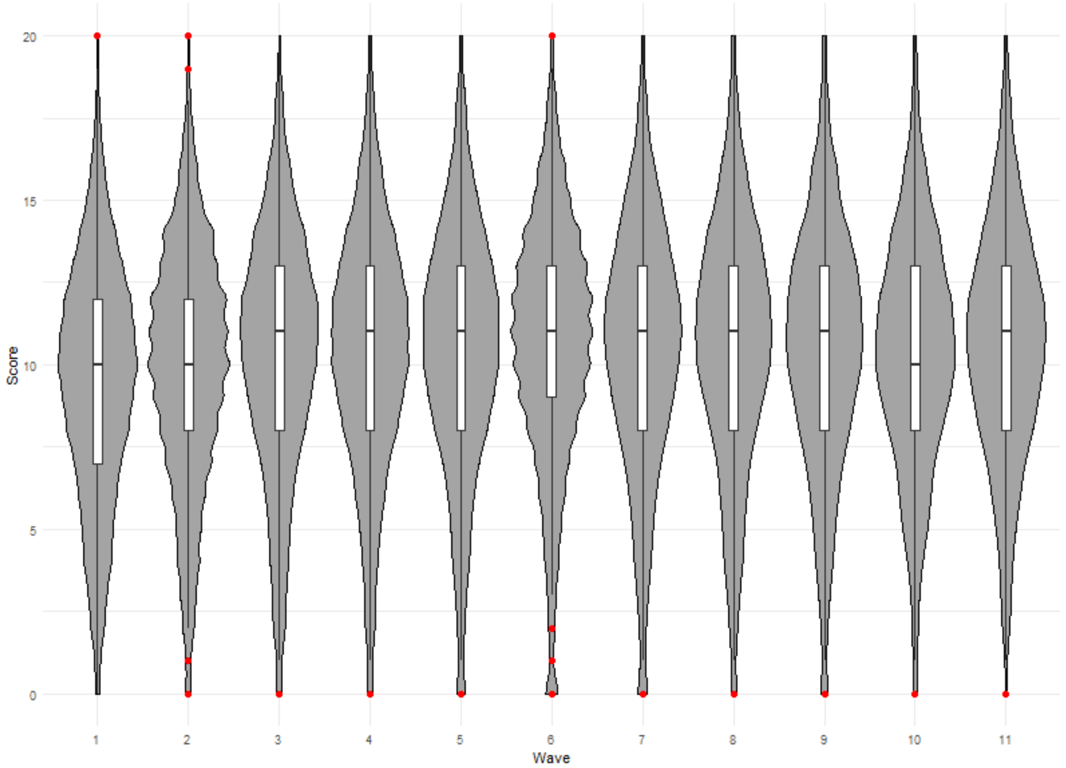


**Figure S1: Distribution of scores on the word recall test (immediate and delayed summed)**

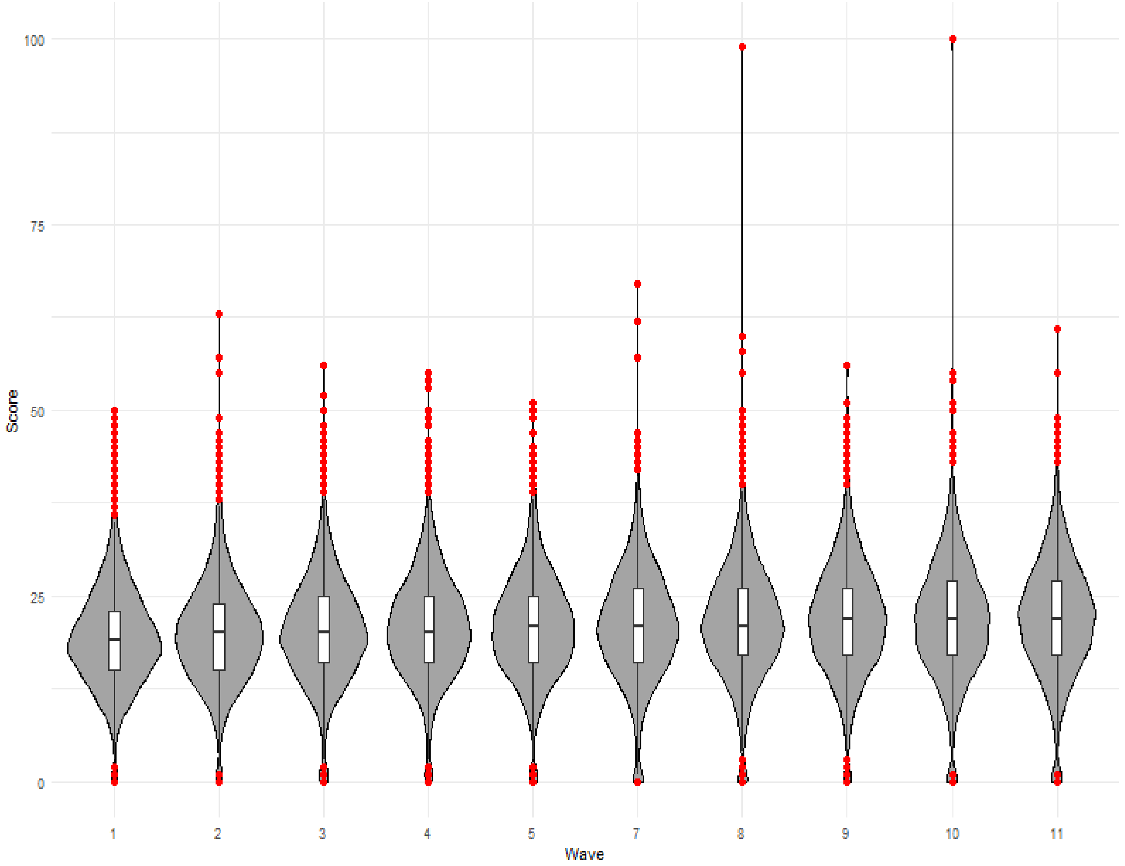


**Figure S2: Distribution of scores on the animal naming test (before outliers were removed).** During data cleaning, we set to NA all the results beyond 4 standard deviations (4SD = 54) as this plot demonstrated they were clear outliers. Scores of 0 were also set to NA since they likely indicate the participant did not complete or understand the task

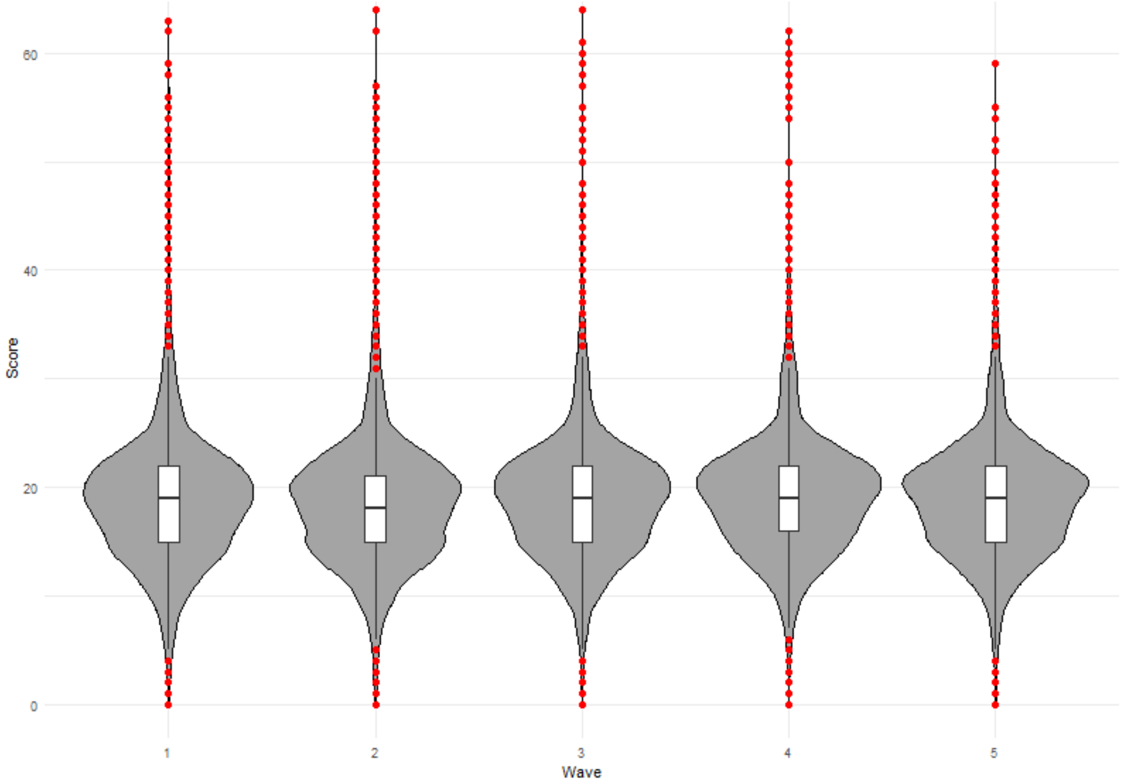


**Figure S3: Distribution of scores on the letter cancellation test.** During data cleaning, we set to NA all the scores reported as 0 since they likely indicate the participant did not complete or understand the task

#### II. Approach to building the factor-of-curves of cognitive function

To use the factor-of-curves approach, individual cognitive tests should first be more highly correlated when measured at the same wave than across waves which was demonstrable in our dataset (see correlation matrix below in Figure S4).

The next step, according to Wickrama et al.,[59] is to build a parallel process model combining all three tests where we allowed the residual variance of the latent trajectory for each cognitive test to correlate with the residual variances of the next and previous wave as well as within each domain across waves. In our model, this significantly improved fit (CFI = 0.966 → 0.983, RMSEA = 0.023 → 0.018, $p$ < 0.001 on the Satorra-Bentler Scaled Chi-Square)[49] and we found stronger correlation among the slopes of the various domains rather than between the intercept and slope of each individual domain. This indicates that there is likely a global, second order growth factor we could identify, and allows us to build a factor-of-curves model using these data.

The final higher-order model built from this parallel process and used in the present study allowed the within-wave covariance of each cognitive test (at each wave, all three tests were correlated with each other); however, the test-specific covariation of intercept and slope was not supported by the data and led to a Heywood case (i.e., an impossible parameter estimate) so to allow for model convergence within boundaries, the covariance between intercept and slope of each cognitive tests was set to zero.

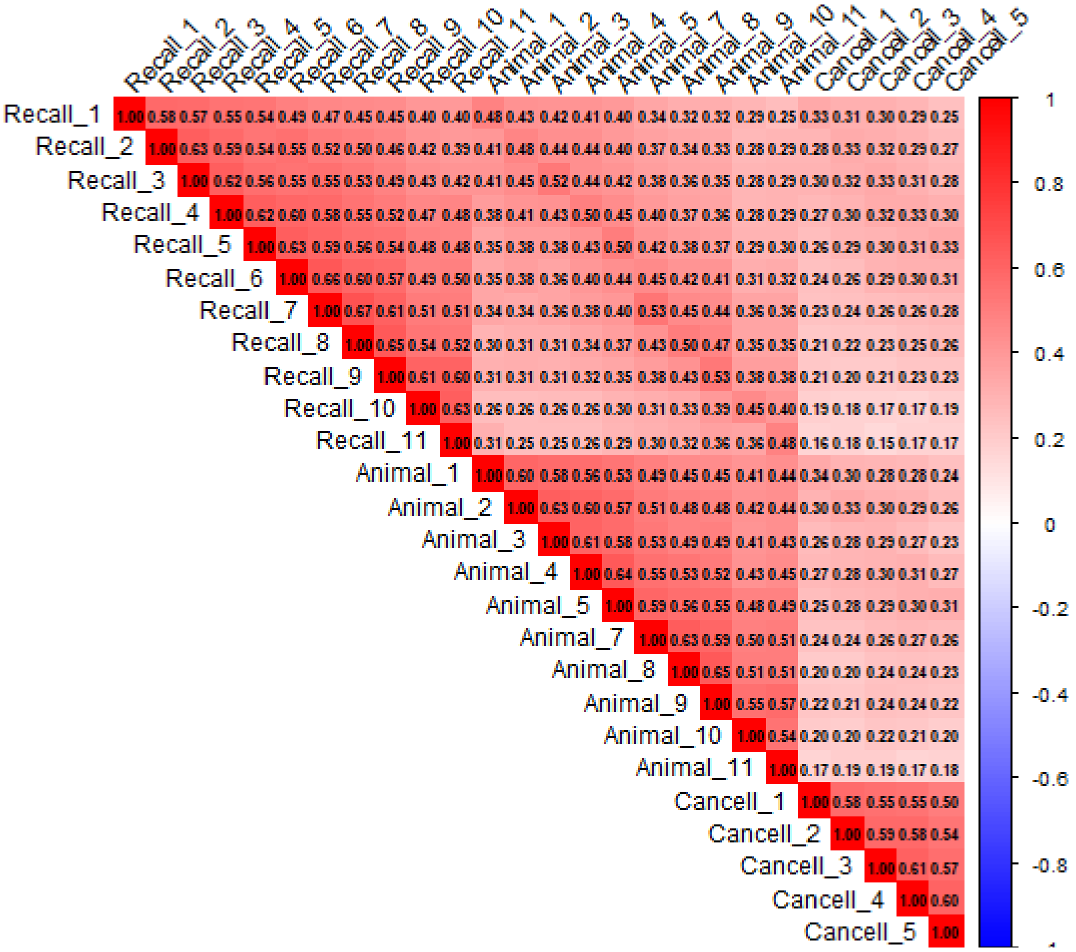


**Figure S4: Correlation matrix of the cognitive tests at all time points.** Animal_# = Animal naming test, Cancell_# = Letter cancellation test. This shows a higher correlation between each test among the same wave of data collection than across waves.

#### III. Patterns of missingness

On average there was 63% missingness on all 4 dependent variables across all waves, however, when examining the dataset wave by wave – ignoring missing data by design - this fell to 6% on average (see figures below).

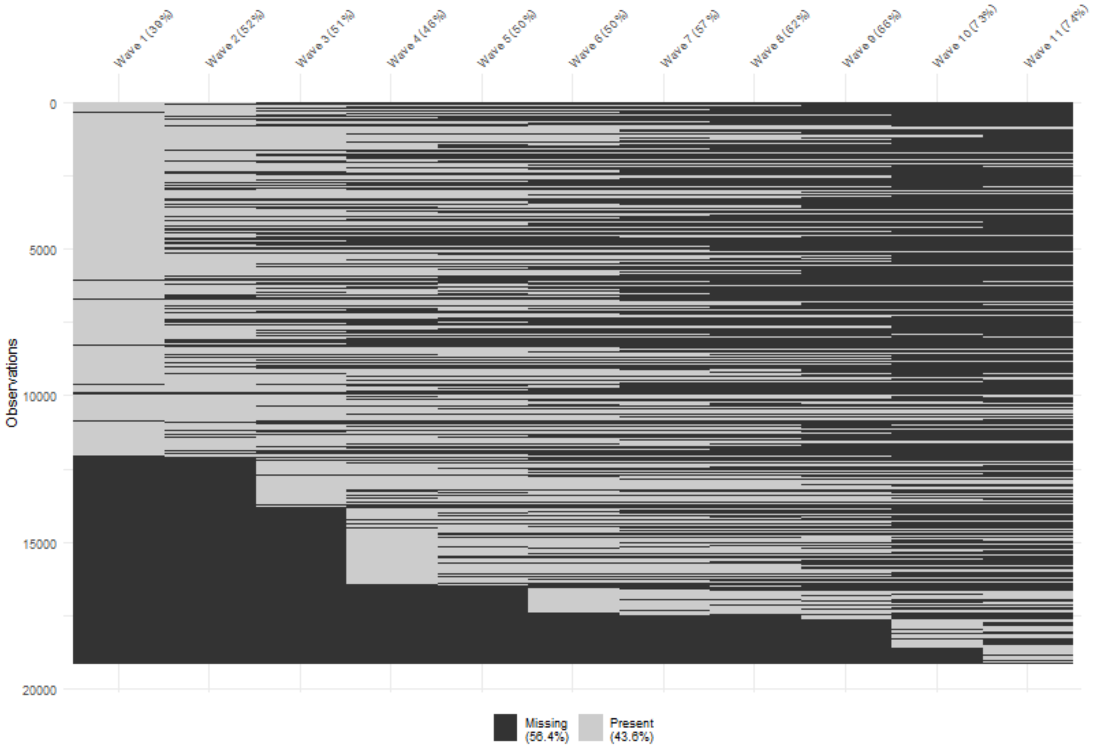


**Figure S5: Patterns of missingness in the recall variable**

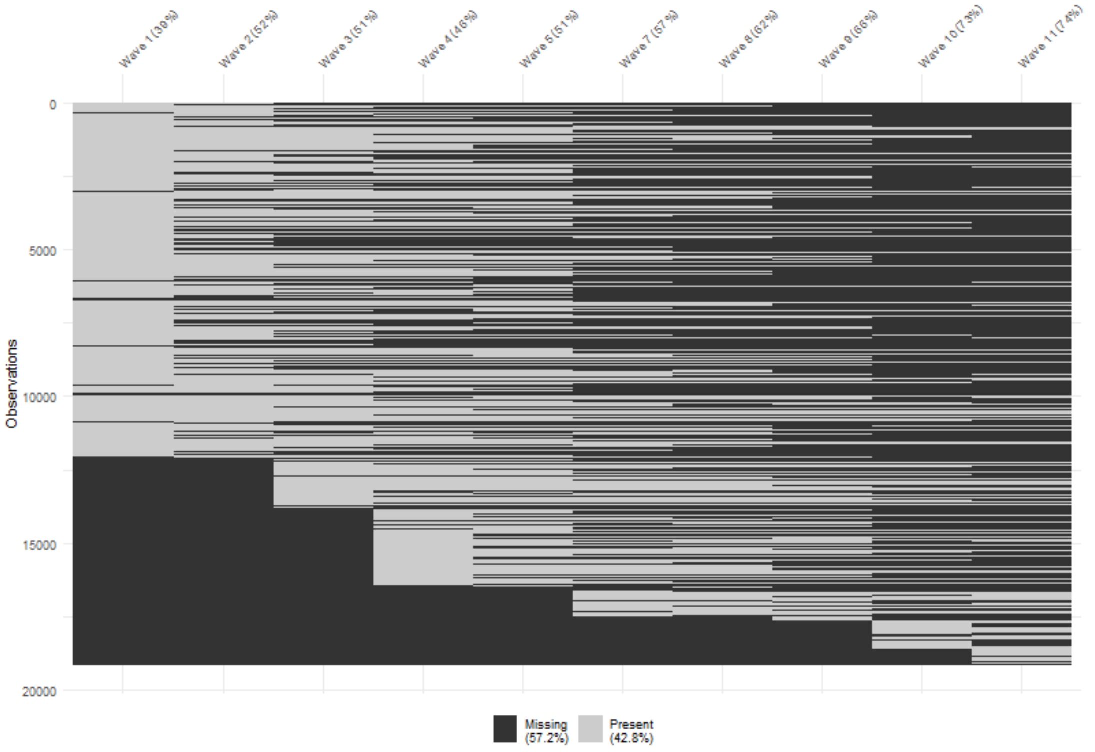


**Figure S6: Patterns of missingness in the animal naming variable**

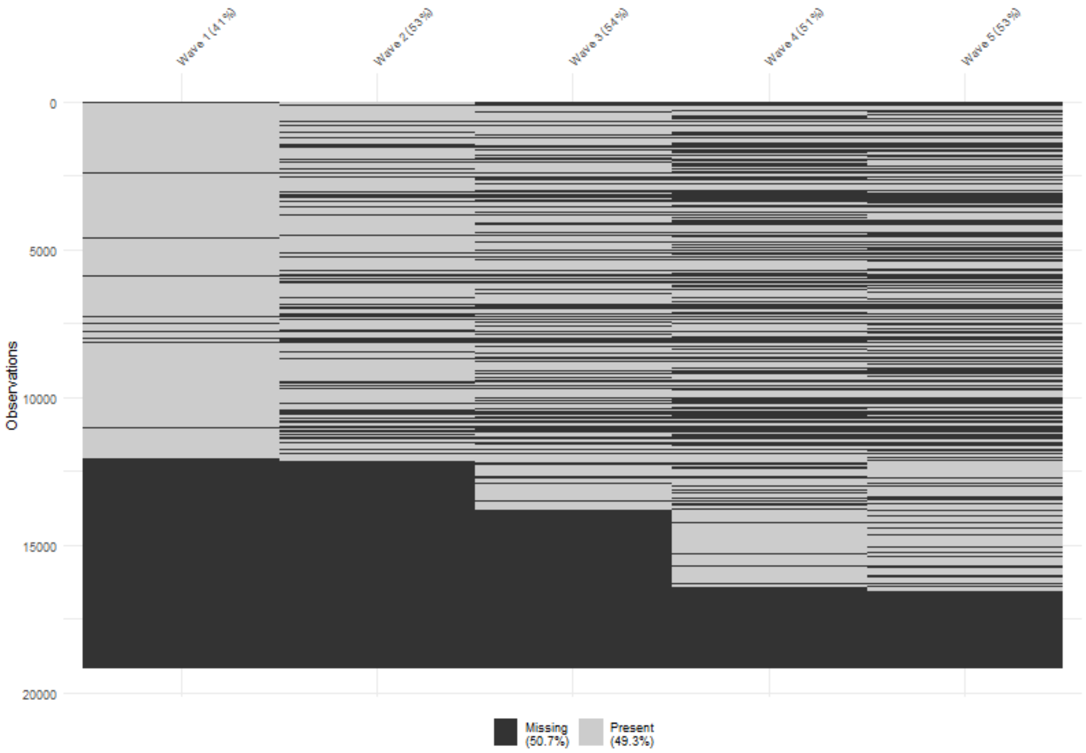


**Figure S7: Patterns of missingness in the letter cancellation variable**

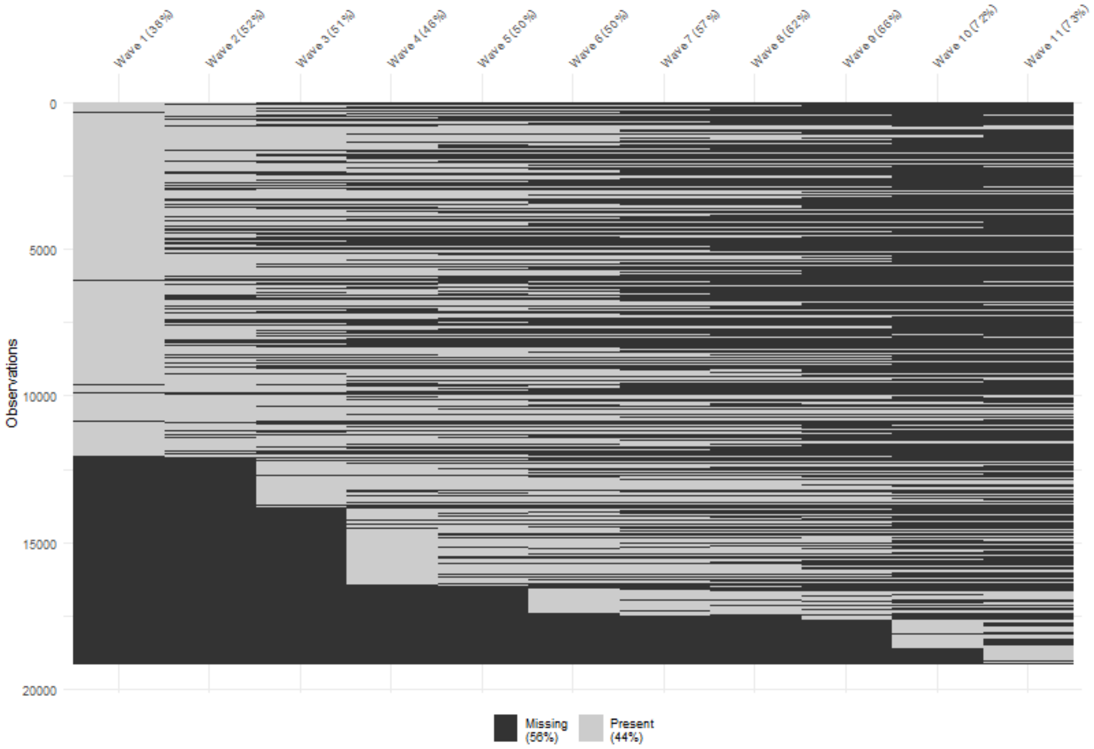


**Figure S8: Patterns of missingness in the pain variable**

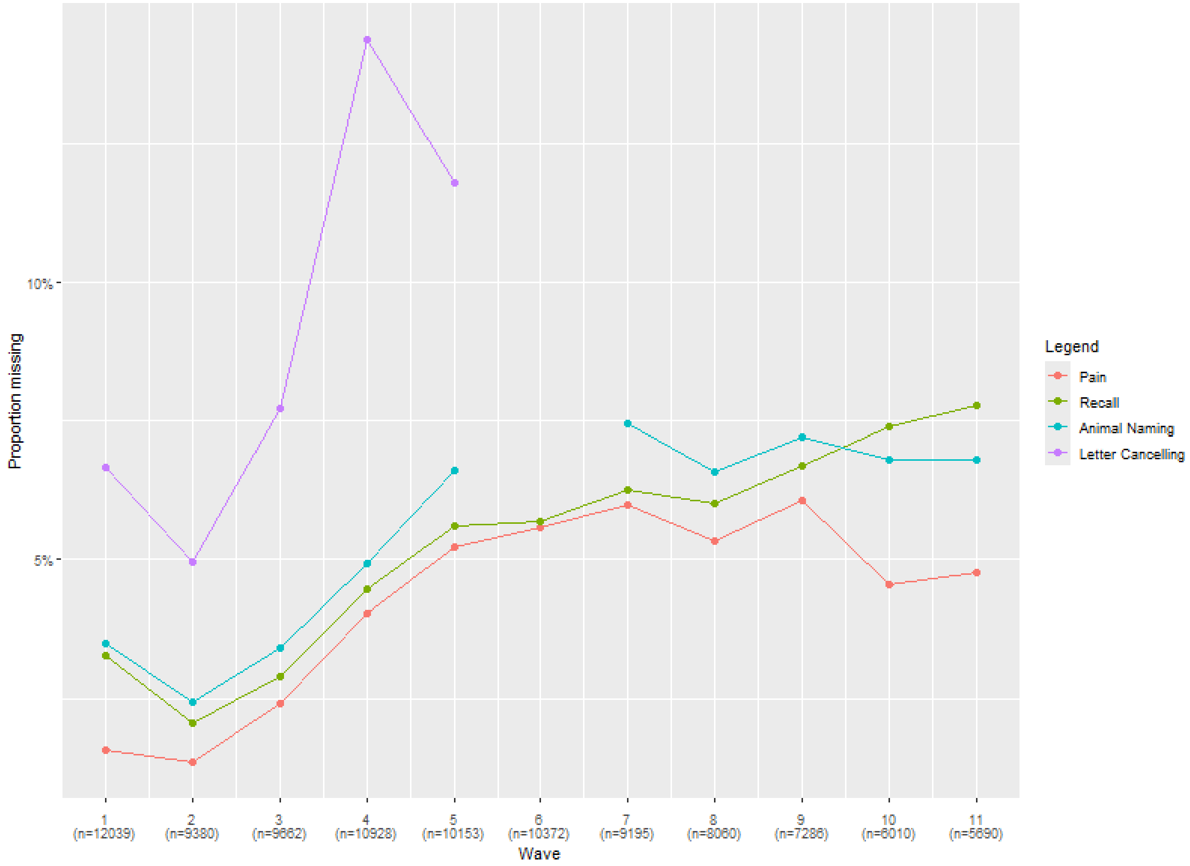


**Figure S9. Proportion of missing data wave by wave**

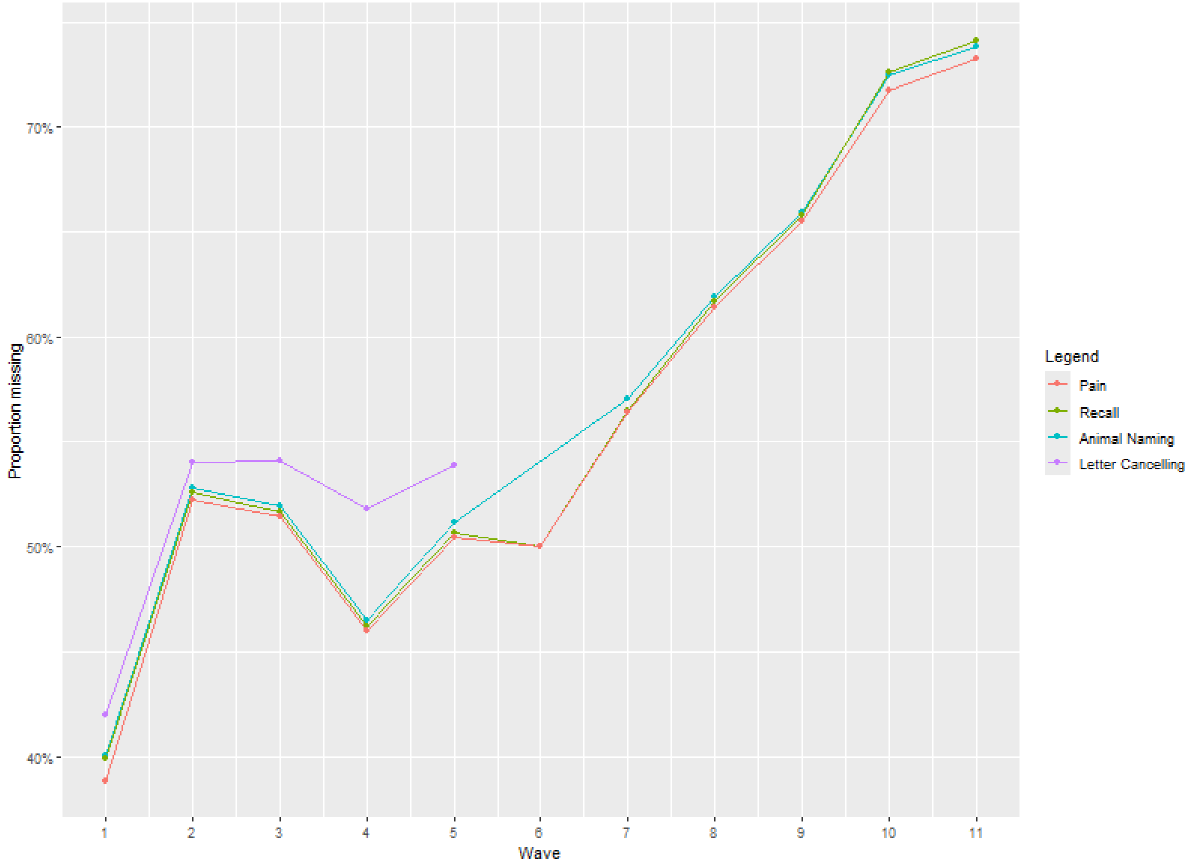


**Figure S10: Proportion of missing data on full dataset (N = 19,376)**

| | W2 | W3 | W4 | W5 | W6 | W7 | W8 | W9 | W10 | W11 | Mean |
|---|---|---|---|---|---|---|---|---|---|---|---|
| 0 | 59.82 | 58.36 | 58.85 | 59.50 | 54.38 | 55.44 | 57.61 | 53.23 | 53.55 | 52.99 | 56.37 |
| 1 | 9.90 | 10.06 | 10.93 | 10.59 | 12.20 | 12.81 | 11.22 | 12.31 | 11.04 | 11.49 | 11.25 |
| 2 | 18.32 | 22.20 | 19.83 | 19.77 | 22.11 | 21.88 | 20.26 | 21.48 | 25.15 | 25.40 | 21.64 |
| 3 | 11.96 | 9.38 | 10.39 | 10.14 | 11.30 | 9.88 | 10.92 | 12.99 | 10.25 | 10.13 | 10.73 |

**Table S1: Rate of attrition at each wave of data collection for the pain variable in %.** Categories 0, 1, 2, 3 represent the response to the pain variable (none, mild, moderate, severe) at the previous wave.

#### IV. Higher order factor loadings

| Variable | | Factor-of-curves LGCM | | | |
|---|---|---|---|---|---|
| | | Estimate | SE | z-score | p value |
| **Loading on intercept of G** | Recall | 0.896 | 0.007 | 121.895 | <0.001 |
| | Animal | 0.828 | 0.009 | 94.507 | <0.001 |
| | Cancell | 0.665 | 0.008 | 79.931 | <0.001 |
| **Loading on slope of G** | Recall | 0.835 | 0.067 | 12.404 | <0.001 |
| | Animal | 0.982 | 0.075 | 13.163 | <0.001 |
| | Cancell | 0.633 | 0.093 | 6.794 | <0.001 |
| | | Model 1 (No covariates) | | | |
| | | Estimate | SE | z-score | p-value |
| **Loading on intercept of G** | Recall | 0.895 | 0.007 | 130.265 | <0.001 |
| | Animal | 0.831 | 0.008 | 103.367 | <0.001 |
| | Cancell | 0.664 | 0.008 | 80.06 | <0.001 |
| **Loading on slope of G** | Recall | 0.851 | 0.053 | 15.959 | <0.001 |
| | Animal | 0.962 | 0.057 | 16.839 | <0.001 |
| | Cancell | 0.632 | 0.091 | 6.946 | <0.001 |
| | | Model 2 (Age and sex) | | | |
| | | Estimate | SE | z-score | p value |
| **Loading on intercept of G** | Recall | 0.925 | 0.006 | 161.867 | <0.001 |
| | Animal | 0.787 | 0.007 | 109.179 | <0.001 |
| | Cancell | 0.669 | 0.009 | 74.445 | <0.001 |
| **Loading on slope of G** | Recall | 0.909 | 0.019 | 46.971 | <0.001 |
| | Animal | 0.899 | 0.021 | 42.433 | <0.001 |
| | Cancell | 0.725 | 0.091 | 7.943 | <0.001 |
| | | Model 3 (Age, sex, ethnicity, socio-economic status and comorbidities) | | | |
| | | Estimate | SE | z-score | p-value |
| **Loading on intercept of G** | Recall | 0.916 | 0.005 | 173.553 | <0.001 |
| | Animal | 0.802 | 0.007 | 107.407 | <0.001 |
| | Cancell | 0.669 | 0.009 | 78.477 | <0.001 |
| **Loading on slope of G** | Recall | 0.914 | 0.023 | 39.614 | <0.001 |
| | Animal | 0.885 | 0.026 | 34.591 | <0.001 |
| | Cancell | 0.73 | 0.091 | 8.026 | <0.001 |

**Table S2: Loadings on higher order factor from each cognitive test in the LGCM (before adding pain to the model) and 3 parallel process models.** G = General cognitive function higher order factor, Recall = Word Recall test, Animal = Animal naming test, Cancell = Letter cancellation test, SE = Standard error. P-values have not undergone FDR correction.

### V. Path diagram of the parallel process of pain on individual cognitive test

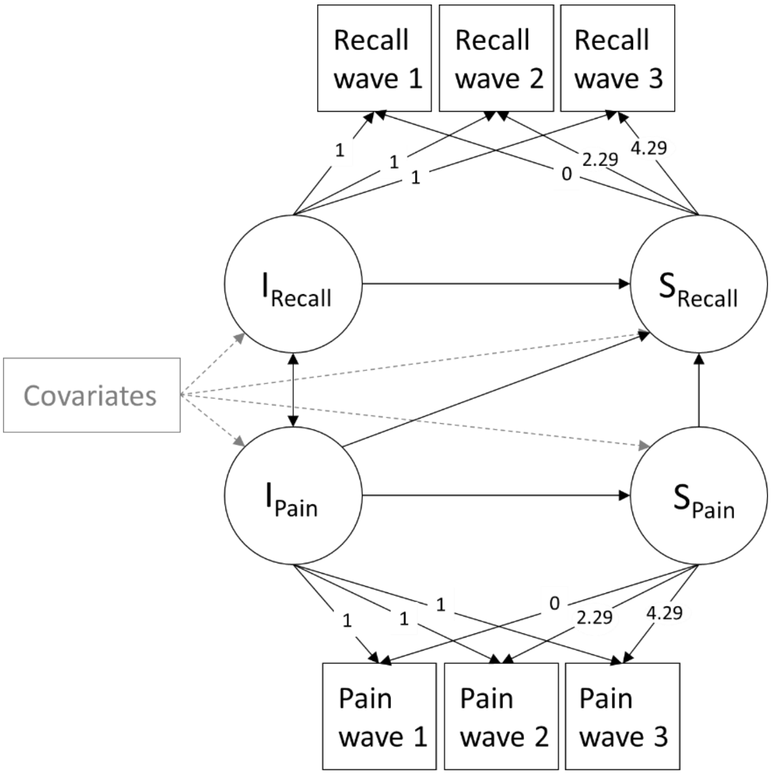


**Figure 1S11: Simplified path diagram of the parallel process latent growth curve model of pain severity and a single cognitive test (word recall for illustrative purposes).** Circles are latent factors, squares represent observed variables, single-headed arrows are regression paths and double-headed arrows are correlation paths. Dashed grey lines are regression paths for covariates. I = Intercept, S = Slope, Recall = Word recall test. For illustrative purposes, only the first three waves are shown, but all 11 waves were used in the model.

### VI. Descriptive statistics on covariates

| Wave number | 1 | 2 | 3 | 4 | 5 | 6 | 7 | 8 | 9 | 10 | 11 |
|---|---|---|---|---|---|---|---|---|---|---|---|
| Years since wave 1 | 0 | 2.29 | 4.29 | 6.25 | 8.29 | 10.21 | 12.21 | 14.21 | 16.29 | 19.79 | 21.59 |
| N | 12039 | 9380 | 9662 | 10928 | 10153 | 10372 | 9195 | 8060 | 7286 | 6010 | 5690 |
| **Sex (Female)** | | | | | | | | | | | |
| N | 6716 | 5267 | 5398 | 6033 | 5616 | 5693 | 5065 | 4448 | 4013 | 3230 | 3083 |
| % | 55.8 | 56.2 | 55.9 | 55.2 | 55.3 | 54.9 | 55.1 | 55.2 | 55.1 | 53.7 | 54.2 |
| **Age** | | | | | | | | | | | |
| N | 12037 | 9380 | 9662 | 10928 | 10153 | 10372 | 9195 | 8060 | 7286 | 6010 | 5550 |
| Mean | 64.27 | 65.86 | 64.74 | 65.41 | 66.97 | 66.97 | 68.28 | 69.75 | 71.02 | 71.50 | 72.15 |
| SD | 10.78 | 10.31 | 10.92 | 10.01 | 9.63 | 9.83 | 9.47 | 8.92 | 8.62 | 8.04 | 7.21 |
| **Ethnicity (White)** | | | | | | | | | | | |
| N | 11601 | 9145 | 9365 | 10553 | 9808 | 9979 | 8846 | 7774 | 7015 | 5713 | 5342 |
| % | 98.4 | 97.6 | 97.0 | 96.7 | 96.6 | 96.2 | 96.2 | 96.5 | 96.3 | 95.4 | 94.2 |
| **Socioeconomic Status** | | | | | | | | | | | |
| SES 1 | | | | | | | | | | | |
| N | 1042 | 834 | 907 | 1041 | 1019 | 1096 | 964 | 845 | 789 | 773 | 869 |
| % | 8.8 | 9.0 | 9.5 | 9.9 | 10.3 | 10.8 | 10.9 | 10.8 | 11.3 | 13.5 | 15.6 |
| SES 2 | | | | | | | | | | | |
| N | 2077 | 2019 | 2164 | 2490 | 2365 | 2436 | 2128 | 1897 | 1710 | 1519 | 1522 |
| % | 17.6 | 21.7 | 22.6 | 23.7 | 23.9 | 24.1 | 24.1 | 24.3 | 24.5 | 26.5 | 27.4 |
| SES 3 | | | | | | | | | | | |
| N | 2067 | 1299 | 1330 | 1430 | 1366 | 1377 | 1252 | 1113 | 992 | 812 | 819 |
| % | 17.5 | 13.9 | 13.9 | 13.6 | 13.8 | 13.6 | 14.2 | 14.3 | 14.2 | 14.2 | 14.7 |
| SES 4 | | | | | | | | | | | |
| N | 1184 | 978 | 1069 | 1223 | 1157 | 1187 | 1099 | 967 | 885 | 723 | 568 |
| % | 10.1 | 10.5 | 11.2 | 11.6 | 11.7 | 11.7 | 12.4 | 12.4 | 12.7 | 12.6 | 10.2 |
| SES 5 | | | | | | | | | | | |
| N | 1361 | 994 | 1003 | 1019 | 954 | 928 | 776 | 659 | 584 | 393 | 336 |
| % | 11.6 | 10.7 | 10.5 | 9.7 | 9.6 | 9.2 | 8.8 | 8.4 | 8.4 | 6.9 | 6.0 |
| SES 6 | | | | | | | | | | | |
| N | 2153 | 1667 | 1642 | 1805 | 1652 | 1695 | 1458 | 1303 | 1132 | 838 | 697 |
| % | 18.3 | 17.9 | 17.2 | 17.2 | 16.7 | 16.8 | 16.5 | 16.7 | 16.2 | 14.6 | 12.5 |
| SES 7 | | | | | | | | | | | |
| N | 1897 | 1381 | 1314 | 1371 | 1288 | 1300 | 1086 | 956 | 826 | 616 | 602 |
| % | 16.1 | 14.8 | 13.8 | 13.1 | 13.0 | 12.9 | 12.3 | 12.3 | 11.9 | 10.8 | 10.8 |
| SES 8 | | | | | | | | | | | |
| N | 0 | 144 | 126 | 120 | 99 | 94 | 76 | 60 | 50 | 48 | 150 |
| % | 0.0 | 1.5 | 1.3 | 1.1 | 1.0 | 0.9 | 0.9 | 0.8 | 0.7 | 0.8 | 2.7 |
| **Comorbidities*** | | | | | | | | | | | |
| Diabetes | | | | | | | | | | | |
| N | 864 | 383 | 255 | 377 | 288 | 308 | 259 | 193 | 263 | 819 | 246 |
| % | 7.2 | 4.1 | 2.6 | 3.5 | 2.8 | 3.0 | 2.8 | 2.4 | 3.6 | 13.6 | 4.3 |
| Hypertension | | | | | | | | | | | |
| N | 4465 | 1704 | 914 | 1282 | 772 | 802 | 593 | 469 | 489 | 2874 | 755 |
| % | 37.1 | 18.2 | 9.5 | 11.7 | 7.6 | 7.7 | 6.5 | 5.8 | 6.7 | 47.9 | 13.3 |
| Heart disease | | | | | | | | | | | |
| N | 2156 | 851 | 491 | 726 | 617 | 611 | 667 | 576 | 632 | 1522 | 507 |
| % | 17.9 | 9.1 | 5.1 | 6.7 | 6.1 | 5.9 | 7.3 | 7.2 | 8.7 | 25.3 | 8.9 |
| Lung disease | | | | | | | | | | | |
| N | 762 | 687 | 129 | 244 | 140 | 179 | 145 | 114 | 107 | 434 | 138 |
| % | 6.3 | 7.3 | 1.3 | 2.2 | 1.4 | 1.7 | 1.6 | 1.4 | 1.5 | 7.2 | 2.4 |
| Stroke | | | | | | | | | | | |
| N | 511 | 166 | 107 | 176 | 148 | 154 | 134 | 148 | 105 | 363 | 94 |
| % | 4.2 | 1.8 | 1.1 | 1.6 | 1.5 | 1.5 | 1.5 | 1.8 | 1.4 | 6.0 | 1.7 |
| Cancer | | | | | | | | | | | |

| | | | | | | | | | | | |
|---|---|---|---|---|---|---|---|---|---|---|---|
| N | 727 | 703 | 233 | 346 | 304 | 303 | 279 | 268 | 242 | 981 | 233 |
| % | 6.0 | 7.5 | 2.4 | 3.2 | 3.0 | 2.9 | 3.0 | 3.3 | 3.3 | 16.3 | 4.1 |

**Table S3: Descriptive statistics for covariates across all 11 waves.** SES = Socioeconomic Status based on the National Statistics Socio-Economic Classification.[25] *At wave 10, the question "Diagnosed [condition] newly reported" changed to "Whether ever been told had [condition] by doctor". This had no impact on our analysis since we adjusted the third model for comorbidities in a time-invariant fashion and considered the participants as having a condition if they ever reported it, at any wave of the dataset.

#### VII. Individual and mean trajectories for each cognitive test

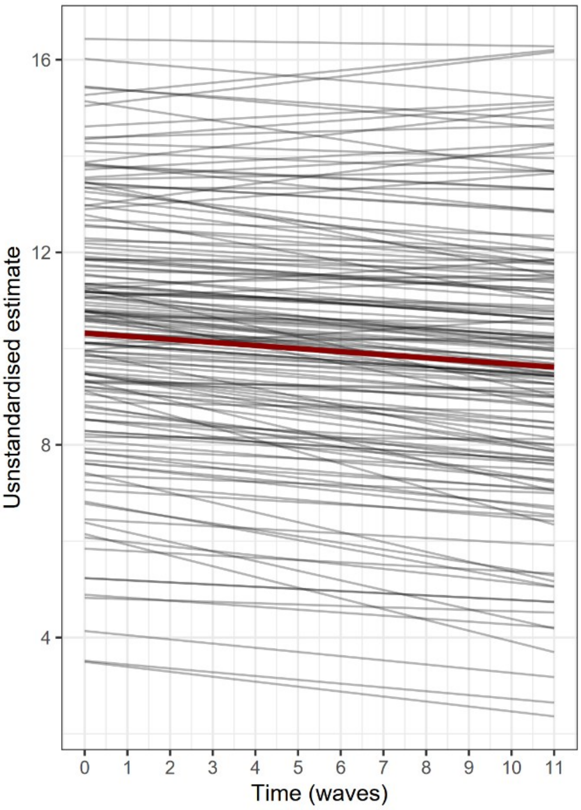


**Figure S12: Trajectories of recall test results.** The unstandardised estimates of the word recall test intercept and slope from a random sample of 150 participants estimated by measurement model LGCM are shown as well as the mean unstandardised estimate from the model in dark red (intercept β = 10.320 and slope β = -0.064)

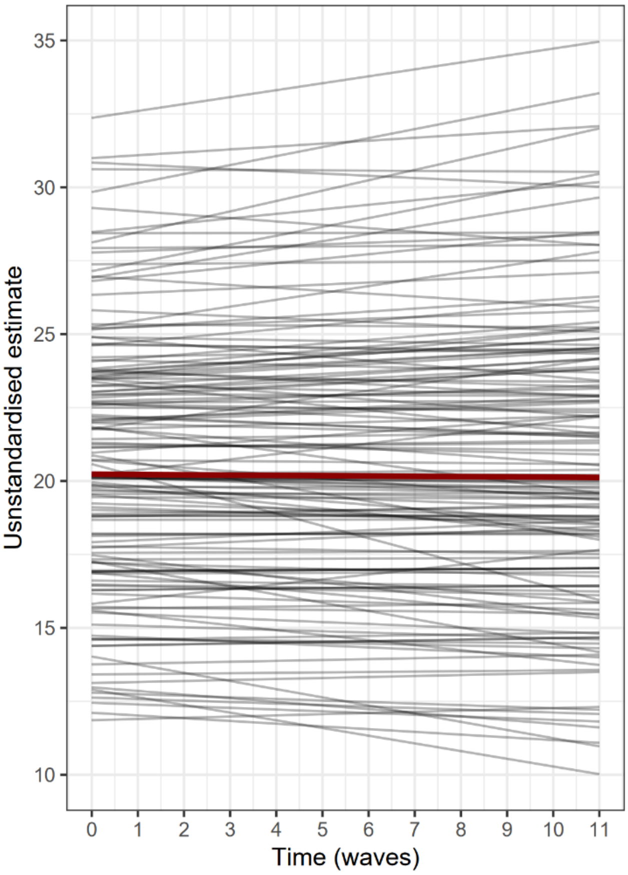


**Figure S13 Trajectories of animal naming test results.** The unstandardised estimates of the animal naming test intercept and slope from a random sample of 150 participants estimated by measurement model LGCM are shown as well as the mean unstandardised estimate from the model in dark red (intercept β = 20.231 and slope β = -0.010)

**Figure S14: Trajectories of letter cancellation test results.** The unstandardised estimates of letter cancellation test results intercept and slope from a random sample of 150 participants estimated by measurement model LGCM are shown as well as the mean unstandardised estimate from the model in dark red (intercept β = 19.012 and slope β = -0.088)

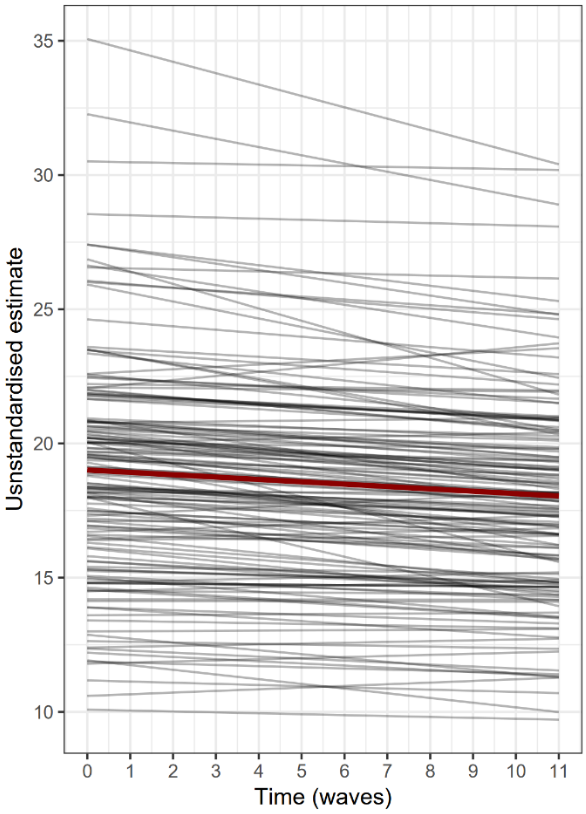


#### VIII. Rank order stability analysis

Rank-order stability was investigated for the four observed dependent variables (pain and three cognitive tests) and found to be high on all outcomes. To allow equal-length comparisons across measures we focussed on rank-order stability over waves 1–5, 5–10 and 1-10 and letter cancellation is reported on 1-3, 3-5 and 1-5 due to the lack of measurement beyond wave 5. The model-implied correlations between latent true scores during the first half of the study (at waves 1 to 5) were large: pain r = 0.934, recall r = 0.945 and animal naming r = 0.938. Stability over the second half (at waves 5 to 10) remained high: pain r = 0.873, recall r = 0.929, animal naming r = 0.918. Finally, overall rank order from waves 1 to 10 was still very stable: pain r = 0.641, recall r = 0.755 and animal naming r = 0.723. A similar pattern was found for the letter cancellation test: waves 1 to 3 r = 0.966, waves 3 to 5 r = 0.969 and waves 1 to 5 r = 0.874. These results are in concordance with the high variance in intercept and low variance in slopes for all four observed dependent variables, indicating that while participants differed in their initial levels of reported pain and measured cognitive function, their rates of change are similar, often maintaining the order in which they began the study.

### IX. Regression coefficients on covariates

| Variable | | Model 2 (Age and sex) | | | | Model 3 (Age, sex, ethnicity, socio-economic status and comorbidities) | | | |
|---|---|---|---|---|---|---|---|---|---|
| | | Estimate | SE | z-score | p value | Estimate | SE | z-score | p-value |
| **Intercept of G regressed on** | Age | -0.598 | 0.008 | -75.956 | <0.001 | -0.562 | 0.008 | -70.040 | <0.001 |
| | Sex (Female) | 0.086 | 0.010 | 8.930 | <0.001 | 0.113 | 0.010 | 11.632 | <0.001 |
| | Ethnicity (White) | | | | | 0.168 | 0.011 | 14.986 | <0.001 |
| | SES | | | | | -0.339 | 0.008 | -44.457 | <0.001 |
| | Diabetes | | | | | -0.013 | 0.008 | -1.715 | 0.086 |
| | Hypertension | | | | | -0.046 | 0.008 | -5.713 | <0.001 |
| | Cancer | | | | | 0.029 | 0.007 | 3.830 | <0.001 |
| | Heart disease | | | | | 0.006 | 0.008 | 0.766 | 0.443 |
| | Lung disease | | | | | -0.022 | 0.008 | -2.903 | 0.004 |
| | Stroke | | | | | -0.044 | 0.008 | -5.212 | <0.001 |
| **Slope of G regressed on** | Age | -0.617 | 0.019 | -33.021 | <0.001 | -0.646 | 0.019 | -34.469 | <0.001 |
| | Sex (Female) | 0.025 | 0.020 | 1.253 | 0.210 | 0.034 | 0.022 | 1.549 | 0.121 |
| | Ethnicity (White) | | | | | 0.050 | 0.017 | 2.856 | 0.004 |
| | SES | | | | | -0.046 | 0.018 | -2.547 | 0.011 |
| | Diabetes | | | | | -0.027 | 0.014 | -1.967 | 0.049 |
| | Hypertension | | | | | 0.001 | 0.015 | 0.087 | 0.931 |
| | Cancer | | | | | -0.002 | 0.014 | -0.179 | 0.858 |
| | Heart disease | | | | | 0.005 | 0.014 | 0.349 | 0.727 |
| | Lung disease | | | | | 0.001 | 0.015 | 0.100 | 0.920 |
| | Stroke | | | | | -0.059 | 0.016 | -3.662 | <0.001 |
| **Intercept of pain regressed on** | Age | 0.116 | 0.011 | 10.888 | <0.001 | 0.080 | 0.011 | 7.336 | <0.001 |
| | Sex (Female) | 0.088 | 0.010 | 8.737 | <0.001 | 0.080 | 0.010 | 7.966 | <0.001 |
| | Ethnicity (White) | | | | | -0.032 | 0.012 | -2.760 | 0.006 |
| | SES | | | | | 0.201 | 0.010 | 20.261 | <0.001 |
| | Diabetes | | | | | 0.063 | 0.010 | 6.362 | <0.001 |
| | Hypertension | | | | | -0.006 | 0.011 | -0.550 | 0.582 |
| | Cancer | | | | | 0.005 | 0.010 | 0.533 | 0.594 |
| | Heart disease | | | | | 0.085 | 0.010 | 8.486 | <0.001 |
| | Lung disease | | | | | 0.078 | 0.010 | 7.914 | <0.001 |
| | Stroke | | | | | 0.007 | 0.010 | 0.679 | 0.497 |
| **Slope of pain regressed on** | Age | 0.092 | 0.020 | 4.586 | <0.001 | 0.076 | 0.020 | 3.780 | <0.001 |
| | Sex (Female) | 0.106 | 0.015 | 6.889 | <0.001 | 0.108 | 0.015 | 7.008 | <0.001 |
| | Ethnicity (White) | | | | | -0.007 | 0.016 | -0.422 | 0.673 |
| | SES | | | | | 0.072 | 0.016 | 4.511 | <0.001 |
| | Diabetes | | | | | 0.010 | 0.016 | 0.617 | 0.537 |
| | Hypertension | | | | | 0.027 | 0.017 | 1.597 | 0.110 |
| | Cancer | | | | | 0.020 | 0.015 | 1.340 | 0.180 |
| | Heart disease | | | | | 0.052 | 0.015 | 3.505 | <0.001 |
| | Lung disease | | | | | 0.036 | 0.016 | 2.217 | 0.027 |
| | Stroke | | | | | 0.055 | 0.017 | 3.274 | 0.001 |

**Table S4: Coefficient of covariates in model 2 and 3.** G = General cognitive function higher-order factor, SES = Socioeconomic status and SE = Standard error. P-values have not undergone FDR correction

### X. Model fit information for the 4 parallel process models

| Model | Covariates | AIC | BIC | Sample-size adjusted BIC |
|---|---|---|---|---|
| Parallel process LGCM of pain and general cognitive function | None | 1410120.278 | 1410795.022 | 1410521.718 |
| | Sex and Age | 1400981.591 | 1401719.007 | 1401420.280 |
| | All | 1386863.319 | 1387849.076 | 1387448.655 |
| Parallel process LGCM of pain and word recall | None | 632387.659 | 632677.956 | 632560.372 |
| | Sex and Age | 624450.573 | 624803.591 | 624660.583 |
| | All | 616124.612 | 616727.019 | 616482.318 |
| Parallel process LGCM of pain and animal naming | None | 684830.888 | 685113.334 | 684998.927 |
| | Sex and Age | 679854.933 | 680200.099 | 680060.269 |
| | All | 671526.027 | 672120.611 | 671879.087 |
| Parallel process LGCM of pain and letter cancellation | None | 455865.962 | 456109.177 | 456010.660 |
| | Sex and Age | 451791.351 | 452097.292 | 451973.352 |
| | All | 446955.809 | 447511.275 | 447285.641 |

**Table S5:** Akaike information criteria (AIC), Bayesian information criteria (BIC) and sample-size adjusted BIC for the 4 main models of analysis derived across 3 levels of covariate adjustment

#### XI. Longitudinal association between pain and individual cognitive tests

Nine models were run: three for the word-recall test, three for the animal naming test and three for the letter cancellation test. Each set of three models followed the structure of the primary analysis with no covariates included in model 1, adjustment for age and sex in model 2 and full adjustment for all covariates in model 3. As before, the primary associations of interest were the regression path from latent slope and latent intercept of pain to latent slope of the cognitive test. Table S6 reports the results for the relationship between pain severity and individual cognitive test intercepts and slopes. Overall, the direction, size, and significance of effects are similar for the word-recall and animal naming tests, compared to the factor-of-curves models, but effects differ on the letter cancellation test.

For letter cancellation, there was a small negative correlation between the intercept of pain and the intercept of letter cancellation in the unadjusted model (M1, Table 3). This correlation was slightly attenuated but remained significant in the fully adjusted model (M3, Table 3). In contrast to general cognitive function, the recall test and the animal naming test, the associations between intercept and slope of pain with slope of letter cancellation were small and non-significant even in the unadjusted model, and remained so with adjustment for covariates (Table 3).

Inspection of the 95% confidence intervals confirmed the equivalence between the results of the word recall and animal naming models and the disparity with the letter cancellation models.

| | Model 1 (no covariates) | | | Model 2 (age and sex) | | | Model 3 (age, sex, ethnicity, socio-economic status and comorbidities) | | |
|---|---|---|---|---|---|---|---|---|---|
| Variables | β | SE | *p* | β | SE | *p* | β | SE | *p* |
| **Word recall** | | | | | | | | | |
| Slope of pain → Slope of recall | -0.099 | 0.029 | 0.001 | -0.065 | 0.027 | 0.018 | -0.033 | 0.027 | 0.328 |
| Intercept of pain → Slope of recall | -0.113 | 0.020 | <0.001 | -0.112 | 0.020 | <0.001 | -0.093 | 0.020 | <0.001 |
| Intercept of recall → Slope of recall | -0.113 | 0.024 | <0.001 | -0.192 | 0.027 | <0.001 | -0.232 | 0.029 | <0.001 |
| Intercept of pain → Slope of pain | -0.414 | 0.023 | <0.001 | -0.433 | 0.023 | <0.001 | -0.462 | 0.025 | <0.001 |
| Intercept of recall ↔ Intercept of pain | -0.231 | 0.011 | <0.001 | -0.211 | 0.012 | <0.001 | -0.134 | 0.013 | <0.001 |
| **Animal naming** | | | | | | | | | |
| Slope of pain → Slope of animal | -0.083 | 0.028 | 0.004 | -0.038 | 0.026 | 0.148 | -0.013 | 0.026 | 0.696 |
| Intercept of pain → Slope of animal | -0.148 | 0.020 | <0.001 | -0.120 | 0.019 | <0.001 | -0.104 | 0.019 | <0.001 |
| Intercept of animal → Slope of animal | -0.168 | 0.021 | <0.001 | -0.208 | 0.022 | <0.001 | -0.228 | 0.024 | <0.001 |
| Intercept of pain → Slope of pain | -0.415 | 0.023 | <0.001 | -0.432 | 0.023 | <0.001 | -0.462 | 0.023 | <0.001 |
| Intercept of animal ↔ Intercept of pain | -0.214 | 0.011 | <0.001 | -0.172 | 0.012 | <0.001 | -0.102 | 0.012 | <0.001 |
| **Letter cancellation** | | | | | | | | | |
| Slope of pain → Slope of cancell | 0.022 | 0.043 | 0.608 | -0.008 | 0.042 | 0.847 | 0.005 | 0.042 | 0.908 |
| Intercept of pain → Slope of cancell | -0.045 | 0.029 | 0.133 | -0.048 | 0.029 | 0.111 | -0.043 | 0.030 | 0.171 |
| Intercept of cancell → Slope of cancell | -0.305 | 0.034 | <0.001 | -0.389 | 0.038 | <0.001 | -0.386 | 0.040 | <0.001 |
| Intercept of pain → Slope of pain | -0.418 | 0.023 | <0.001 | -0.433 | 0.023 | <0.001 | -0.462 | 0.023 | <0.001 |
| Intercept of cancell ↔ Intercept of pain | -0.136 | 0.013 | <0.001 | -0.123 | 0.014 | <0.001 | -0.075 | 0.014 | <0.001 |

**Table S6: Summary of results for individual test associations.** Estimates are all standardised. β = Estimate, SE = standard error, *p* = p-value, recall = word recall test, animal = animal naming test, cancell = letter cancellation test. → represent regression paths, ↔ show correlations. The p-values are adjusted within each model for false discovery rate using the Benjamini–Hochberg procedure with α = 0.05.

### XII. Bivariate models

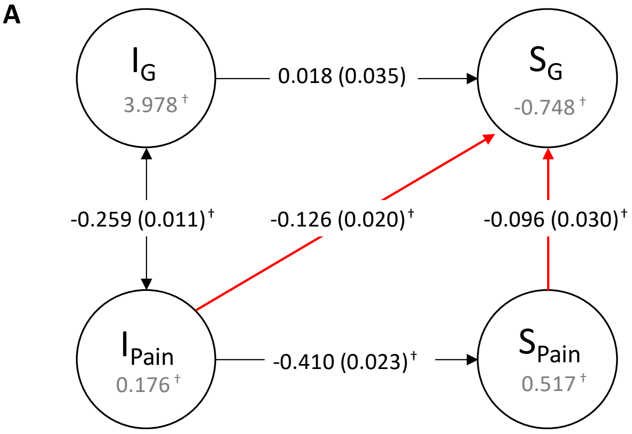


**Figure S15: Side-by-side comparison of original model with bivariate analyses.** A is the original model (model 1 reported in main text); B is running the regression of intercept of pain on slope of cognitive function without the mediating effect of slope of pain; C shows the association between slope of pain and slope of cognitive function without its conditional relationship with intercept of pain

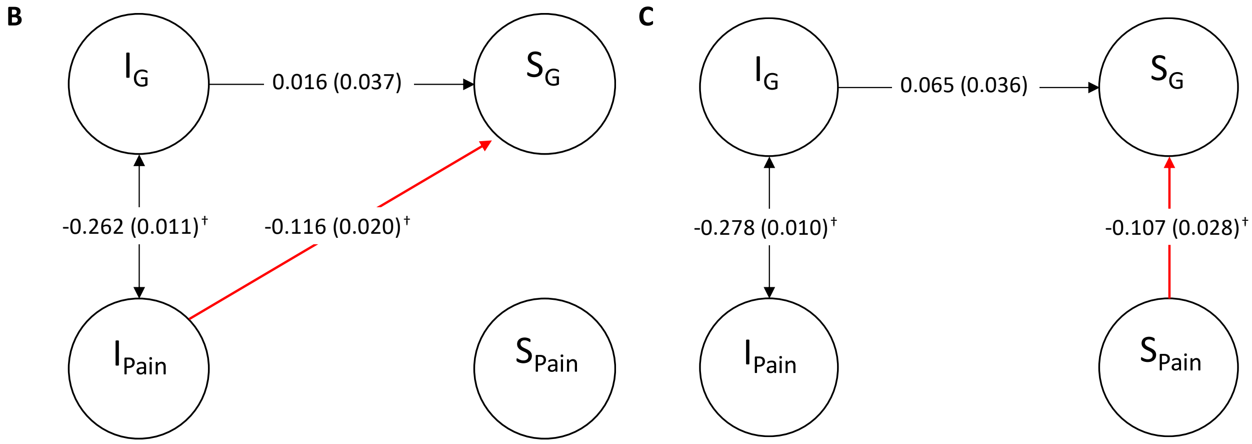

#### XIII. Supplementary sensitivity analyses

We ran the same models as in the primary analysis but without partial invariance of the thresholds. For these three models, MPlus indicates that the third threshold parameter of the pain variable had to be fixed to avoid singularity of the information matrix. We found similar regression and correlation coefficients to the primary analysis models but some discrepancies in the estimated means of intercept and slope of pain severity. The estimates and FDR-adjusted p-values for the three models built under these conditions are in Table S7.

| | Model 1 (no covariates) | | | Model 2 (Age and sex) | | | Model 3 (Age, sex, ethnicity, socio-economic status and comorbidities) | | |
|---|---|---|---|---|---|---|---|---|---|
| Variables | Estimate | SE | *p* | Estimate | SE | *p* | Estimate | SE | *p* |
| **Regressions** | | | | | | | | | |
| Slope of g → Slope of pain | -0.102 | 0.031 | 0.002 | -0.053 | 0.027 | 0.063 | -0.024 | 0.027 | 0.387 |
| Slope of g → Intercept of pain | -0.136 | 0.021 | <0.001 | -0.121 | 0.018 | <0.001 | -0.105 | 0.019 | <0.001 |
| Slope of g → Intercept of g | 0.018 | 0.035 | 0.614 | -0.145 | 0.031 | <0.001 | -0.207 | 0.036 | <0.001 |
| Slope of pain → Intercept of pain | -0.474 | 0.019 | <0.001 | -0.495 | 0.019 | <0.001 | -0.523 | 0.020 | <0.001 |
| **Correlation** | | | | | | | | | |
| Intercept of g ↔ Intercept of pain | -0.256 | 0.011 | <0.001 | -0.238 | 0.012 | <0.001 | -0.154 | 0.013 | <0.001 |
| **Means** | | | | | | | | | |
| Intercept of g | 3.978 | 0.045 | <0.001 | 3.821 | 0.043 | <0.001 | 3.714 | 0.146 | <0.001 |
| Slope of g | -0.648 | 0.204 | 0.002 | -0.459 | 0.133 | 0.001 | -0.289 | 0.022 | 0.170 |
| Intercept of pain | 0.855 | 0.022 | <0.001 | -0.768 | 0.016 | <0.001 | -0.430 | 0.093 | <0.001 |
| Slope of pain | 0.845 | 0.029 | <0.001 | 0.015 | 0.029 | 0.604 | 0.182 | 0.005 | 0.044 |

**Table S7: Models without partial time-invariance of the thresholds:** Estimates are all standardised. SE = Standard Error. g = general cognitive function as determined by the factor-of-curves method. The p-values are adjusted within each model for false discovery rate using the Benjamini–Hochberg procedure with a = 0.05.

Another set of models was run using only waves 1 to 9 to investigate whether the results of the main models are consistent with results using only data collected before the COVID-19 pandemic. This was done because the average interval and the individual-person intervals after wave 9 are much more varied when compared to the relatively consistent intervals between waves 1 to 9. Like the models in the main analysis, these three models' loadings on the first-order factors were set to the mean time (in years) elapsed since wave 1 and the thresholds of the pain variable were partially invariant. The regression and correlation paths were very similar to the primary models. The main difference with previous results lies again with the estimated means of intercept and slope of pain severity. The results of these models are shown in Table S8.

| | Model 1 (no covariates) | | | Model 2 (Age and sex) | | | Model 3 (Age, sex, ethnicity, socio-economic status and comorbidities) | | |
|---|---|---|---|---|---|---|---|---|---|
| Variables | Estimate | SE | *p* | Estimate | SE | *p* | Estimate | SE | *p* |
| **Regressions** | | | | | | | | | |
| Slope of g → Slope of pain | -0.108 | 0.031 | 0.001 | -0.073 | 0.026 | 0.006 | -0.046 | 0.026 | 0.085 |
| Slope of g → Intercept of pain | -0.123 | 0.020 | <0.001 | -0.112 | 0.018 | <0.001 | -0.102 | 0.018 | <0.001 |
| Slope of g → Intercept of g | 0.090 | 0.030 | 0.003 | -0.105 | 0.031 | 0.001 | -0.158 | 0.035 | <0.001 |
| Slope of pain → Intercept of pain | -0.386 | 0.026 | <0.001 | -0.401 | 0.026 | <0.001 | -0.428 | 0.026 | <0.001 |
| **Correlation** | | | | | | | | | |
| Intercept of g ↔ Intercept of pain | -0.253 | 0.011 | <0.001 | -0.235 | 0.012 | <0.001 | -0.145 | 0.014 | <0.001 |
| **Means** | | | | | | | | | |
| Intercept of g | 3.987 | 0.048 | <0.001 | 3.835 | 0.046 | <0.001 | 3.723 | 0.064 | <0.001 |
| Slope of g | -0.751 | 0.150 | <0.001 | -0.561 | 0.124 | <0.001 | 0.321 | 0.199 | 0.107 |
| Intercept of pain | 0.563 | 0.032 | <0.001 | -3.151 | 0.052 | <0.001 | 3.359 | 0.093 | <0.001 |
| Slope of pain | 0.570 | 0.030 | <0.001 | -0.948 | 0.071 | <0.001 | 1.870 | 0.132 | <0.001 |

**Table S8: Models on waves 1-9 (before the COVID-19 pandemic):** Estimates are all standardised. SE = Standard Error. g = general cognitive function as determined by the factor-of-curves method. The p-values are adjusted within each model for false discovery rate using the Benjamini–Hochberg procedure with a = 0.05.
